# Tissue-specific enhancer-gene maps from multimodal single-cell data identify causal disease alleles

**DOI:** 10.1101/2022.10.27.22281574

**Authors:** Saori Sakaue, Kathryn Weinand, Shakson Isaac, Kushal K. Dey, Karthik Jagadeesh, Masahiro Kanai, Gerald F. M. Watts, Zhu Zhu, Accelerating Medicines Partnership® RA/SLE Program and Network, Michael B. Brenner, Andrew McDavid, Laura T. Donlin, Kevin Wei, Alkes L. Price, Soumya Raychaudhuri

## Abstract

Translating genome-wide association study (GWAS) loci into causal variants and genes requires accurate cell-type-specific enhancer-gene maps from disease-relevant tissues. Building enhancer-gene maps is essential but challenging with current experimental methods in primary human tissues. We developed a new non-parametric statistical method, SCENT (Single-Cell ENhancer Target gene mapping) which models association between enhancer chromatin accessibility and gene expression in single-cell multimodal RNA-seq and ATAC-seq data. We applied SCENT to 9 multimodal datasets including > 120,000 single cells and created 23 cell-type-specific enhancer-gene maps. These maps were highly enriched for causal variants in eQTLs and GWAS for 1,143 diseases and traits. We identified likely causal genes for both common and rare diseases. In addition, we were able to link somatic mutation hotspots to target genes. We demonstrate that application of SCENT to multimodal data from disease-relevant human tissue enables the scalable construction of accurate cell-type-specific enhancer-gene maps, essential for defining non-coding variant function.

## Introduction

Genome-wide association studies (GWAS) have comprehensively mapped loci for human diseases^1–4^. These loci harbor untapped insights about causal mechanisms that can point to novel therapeutics^2,5^. However, only rarely are we able to define causal variants or their target genes. Of the hundreds of associated variants in a single locus, only one or a few may be causal; others are associated since they tag causal variants^2,6,7^. Moreover, causal genes are also challenging to determine, since causal variants lie in non-coding regions in 90% of the time^8–10^, may regulate distant genes^11–13^, and may employ context-specific regulatory mechanisms^14–17^.

To define causal variants and genes, previous studies have used both statistical and experimental approaches. Statistical fine-mapping^18–23^ can narrow the set of candidate causal variants, and is more effective when GWAS includes diverse ancestral backgrounds with different allele frequencies and linkage disequilibrium structures (LD)^24–28^. However, these approaches alone are seldom able to identify true causal variants with confidence^7,23, 29–32^. To define causal genes, previous studies have built enhancer-gene maps, that can be used to prioritize causal variants in enhancers and link causal variants to genes they regulate. These maps often require large-scale epigenetic and transcriptomic atlases (e.g., Roadmap^33^, BLUEPRINT^34^, and ENCODE^35^). The enhancer-gene maps have been built from these atlases by correlating epigenetic activity (i.e., enhancer activity; e.g., histone mark ChIP-seq and bulk ATAC-seq) with gene expression (e.g., RNA-seq)^36,37^, by combining epigenetic activity and probability of physical contact with the gene^38,39^, or by integrating multiple linking strategies to create composite scores ^40^. However, current methods largely use bulk tissues or cell lines. Bulk data potentially (i) cannot be easily applied to rare cell populations (ii) obscures the cell-type-specific nature of gene regulation and (iii) requires hundreds of experimentally characterized samples, necessitating consortium-level efforts. While perturbation experiments (e.g., CRISPR interference^41^ or base editing^42^) can point to causal links between enhancers and genes, they are difficult to scale because they require the cell- or tissue-type specific experimental protocols^43^.

Advances in single-cell technologies offer new opportunities for building cell-type specific enhancer-gene maps. Multimodal protocols now enable joint capture of epigenetic activity by ATAC-seq alongside early transcriptional activity with nuclear RNA-seq^44–48^. These methods are easily applied at scale to cells in human primary tissues without disaggregation, enabling query of many samples from disease-relevant tissues. If we establish accurate links between open chromatin enhancers and genes in single cells, statistical power should exceed bulk-tissue-based methods since each observation is at a cell-level resolution. However, the sparse and non-parametric nature of RNA-seq and ATAC-seq in single-cell experiments makes confident identification of these links challenging. To date, most methods use linear regression models to link enhancers and genes (e.g., ArchR^49^ and Signac^50^) despite these features or only utilize co-accessibility of regulatory regions from ATAC-seq but not gene expression from sc-RNA-seq (e.g., Cicero^51^). These previous methods have not generally demonstrated efficacy in practice for causal variant fine-mapping in complex traits.

In this context, we developed <u>S</u>ingle-<u>C</u>ell <u>En</u>hancer <u>T</u>arget gene mapping (SCENT), to accurately map enhancer-gene pairs where an enhancer’s activity (i.e. peak accessibility) is associated with gene expression across individual single cells. We use Poisson regression and non-parametric bootstrapping^52^ to account for the sparsity and non-parametric distributions. We predicted that peaks with gene associations are more likely to be functionally important. We apply SCENT to 9 multimodal datasets to build 23 cell-type specific enhancer-gene maps. We show that SCENT enhancers are highly enriched in statistically fine-mapped likely causal variants for eQTL and GWAS. We use SCENT enhancer-gene map to define causal variants, genes, and cell types in common and rare disease loci and somatic mutation hotspots, which has not been previously demonstrated by conventional enhancer-gene mapping based on bulk-tissues.

## Results

### Overview of SCENT

To identify (1) active *cis*-regulatory regions and (2) their target genes (3) in a given cell type, we leveraged single-cell multimodal datasets. SCENT accurately identifies significant association between chromatin accessibility of regulatory regions (i.e., peaks) from ATAC-seq and gene expression from RNA-seq across individual single cells (**Figure 1a**). Those associations can be used for prioritizing (1) likely causal variants if they are in regulatory regions that are associated with gene expression, (2) likely causal genes if they are associated with the identified regulatory region and (3) the critical cell types based on which map the association is identified in. We assessed whether binarized chromatin accessibility in an ATAC peak is associated with gene expression counts in *cis* (< 500kb from gene body), testing one peak-gene pair at a time in each cell type (see **Methods**). We tested each cell type separately to capture cell-type-specific gene regulation and to avoid spurious peak-gene associations due to gene co-regulation across cell types.

**Figure 1.**
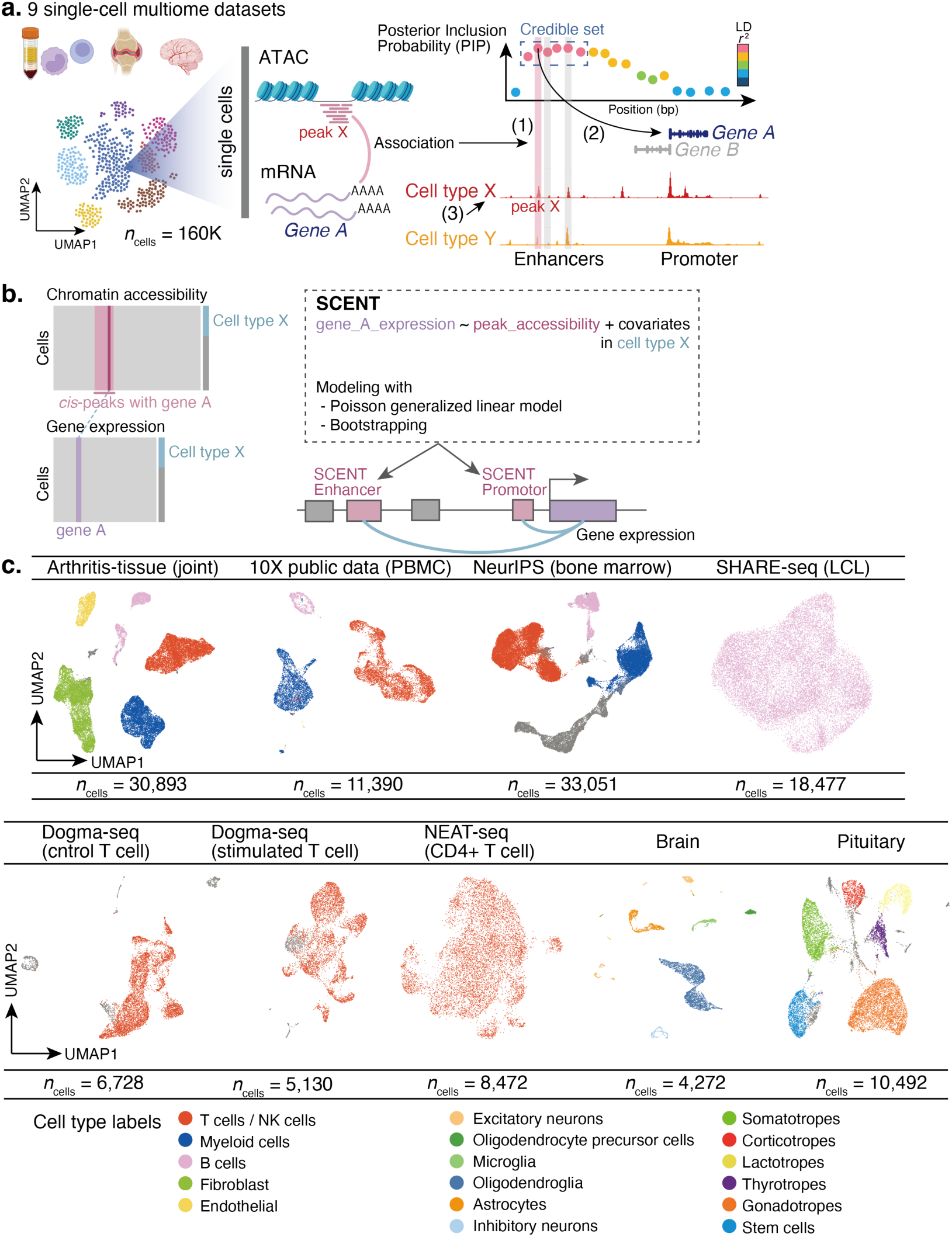
Schematic overview of SCENT and SCENT enhancer-gene pairs across 9 single-cell multimodal datasets. **a**. SCENT identifies (1) active *cis*-regulatory regions and (2) their target genes in (3) a specific cell type. Those SCENT results can be used to define likely causal variants, genes, and cell types for GWAS loci. **b**. SCENT models association between chromatin accessibility from ATAC-seq and gene expression from RNA-seq across individual cells in a given cell type. **c**. 9 single-cell datasets on which we applied SCENT to create 23 cell-type-specific enhancer-gene map. The cells in each dataset are described in UMAP embeddings from RNA-seq and colored by cell types.

Since both RNA-seq and ATAC-seq data are generally sparse^50,53–56^, we used Poisson regression since it was a simple model that has been used effectively for sc-RNA-seq analysis^54,57^:

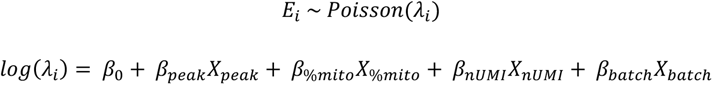

where *E_i_* is the observed expression count of *i*th gene, and *λ_i_* is the expected count under Poisson distribution. *β_peak_* indicates the effect of chromatin accessibility of a peak on *i* th gene expression (see **Methods**) and reflects the strength of the regulatory effect and sign (i.e., enhancing vs. silencing effect). We accounted for donor or batch effects (*X_batch_*) and cell-level technical factors such as percentage of mitochondrial reads (*X_%mito_*).

However, gene expression counts are highly variable across genes (**Figure 1b**; **Supplementary Figure 1a**), and Poisson regression might be suboptimal for highly expressed and dispersed genes. Consequently, we observed inflated statistics when we permuted cell barcodes to disrupt association between ATAC and RNA profiles (**Supplementary Figure 1b**). Common analytical statistical models (e.g., linear, negative binomial and Poisson regression) all demonstrated inflated statistics (**Supplementary Figure 1c-e**). Therefore, to accurately estimate the error and significance of *β_peak_*, we implemented non-parametric bootstrapping framework. Briefly, we resampled cells with replacement from the full data and re-estimated *β′_peak_* up to 50,000 times. We compared this empirical distribution of *β′_peak_* against null hypothesis (*β′_peak_* = 0) to derive the significance of *β_peak_* (i.e., two-sided bootstrapping-based *P* value; see **Methods, Supplementary Figure 2**). The Poisson regression followed by bootstrapping resulted in well-calibrated statistics with appropriate type I error (**Supplementary Figure 1f**).

### Discovery of cell-type-specific SCENT enhancer-gene links

We obtained nine single-cell multimodal datasets from diverse human tissues representing 13 cell-types (immune-related, hematopoietic, neuronal, and pituitary). Since we are interested in autoimmune diseases, we newly generated an inflammatory tissue dataset by obtaining inflamed synovial tissues from ten rheumatoid arthritis (RA) and two osteoarthritis (OA) patients (arthritis-tissue dataset; *n*_donor_ = 12). Applying stringent QC to these multimodal data, we obtained information on 30,893 cells (see **Methods**). In addition, we obtained eight public datasets with 129,672 cells. In total we had data from 160,565 cells^46,58–62^. We analyzed 16,621 genes and 1,193,842 open chromatin peaks in *cis* after QC (4,753,521 peak-gene pairs, 28 median peaks per gene; **Figure 1c**, **Supplementary Figure 3**, Supplementary Table 1). After clustering cells and cell type annotation, we applied SCENT individually to each of the cell types with *n*_cells_ > 500 to construct 23 enhancer-gene maps. SCENT identified 87,648 cell-type-specific peak-gene links (false discovery rate (FDR) < 10%, **Figure 2a**, **Supplementary Figure 4**). Each gene had variable number of associated peaks in *cis* (from 0 to 97, mean = 4.13, **Supplementary Figure 5a**).

**Figure 2.**
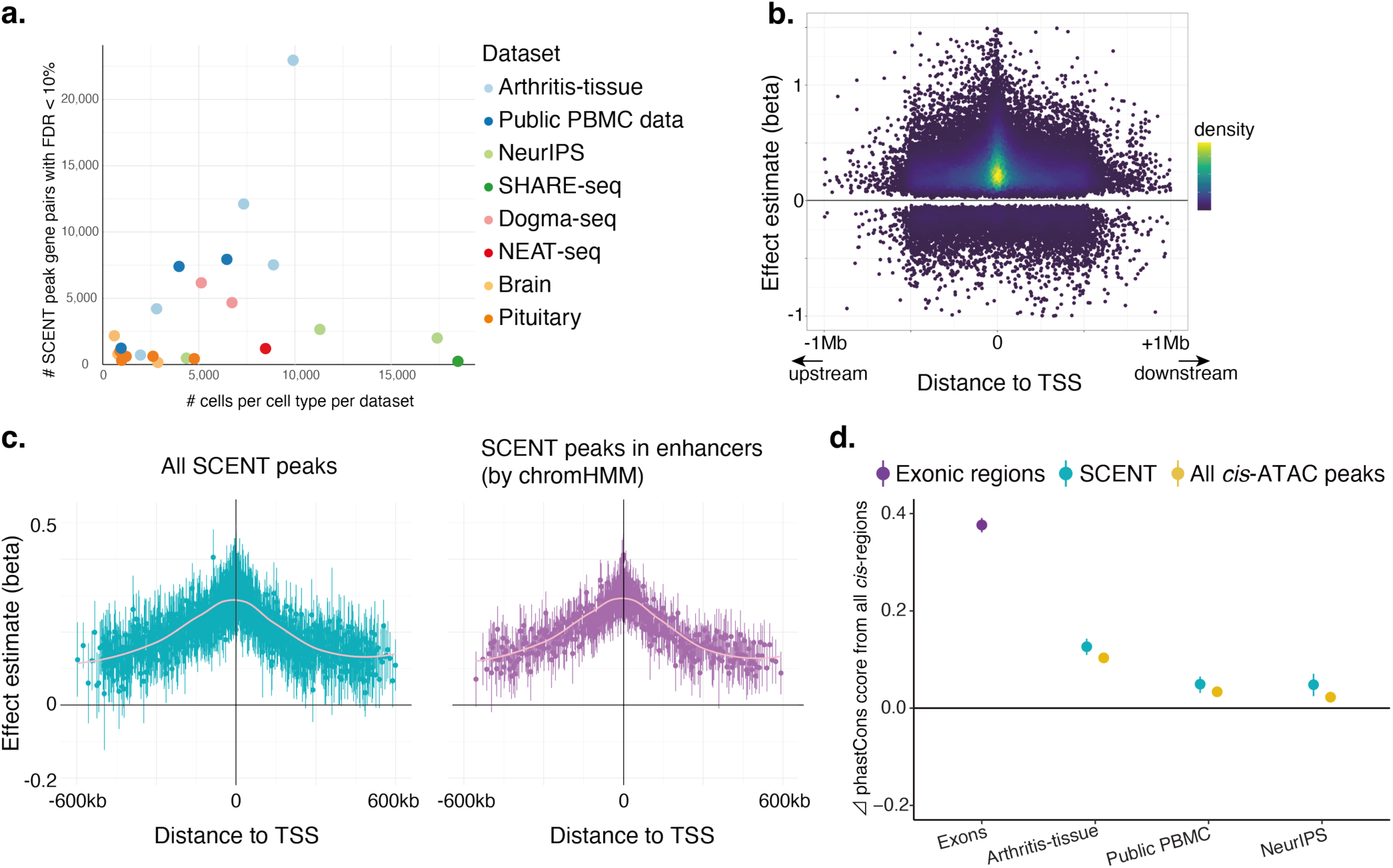
SCENT identified functionally active and evolutionary conserved *cis*-regulatory regions from single-cell multimodal data. **a.** The number of significant gene-peak pairs discovered by SCENT with FDR < 10%. Each dot represents the number of significant gene-peak pairs in a given cell type in a dataset (y-axis) as a function of the number of cells in each cell type in a dataset (x-axis), colored by the dataset. **b.** The effect size (beta) of chromatin accessibility on the gene expression from Poisson regression (y-axis). Each dot is a significant gene-peak pair and plotted against the distance between the peak and the transcription start site (TSS) of the gene, colored as a density plot. **c.** The mean effect size (beta) of chromatin accessibility on the gene expression in arthritis-tissue dataset within each bin of TSS distance. Left; all significant gene-peak links. Right; SCENT peaks within enhancers identified using chromHMM in immune-related tissues. **d**. Mean phastCons score difference (Δ phastCons score) between each annotated region and all *cis*-regulatory non-coding regions. We show the Δ phastCons score for exonic regions (purple) as a reference, and for SCENT (green) and all *cis*-ATAC peaks (yellow) enhancers in each multimodal dataset.

To assess replicability of SCENT peak-gene links, we compared the effects from the arthritis-tissue dataset (discovery; which had the largest number of significant peak-gene pairs) with those from other datasets in the same cell-type (replication) in B cells, T/NK cells and myeloid cells (Supplementary Table 2a). Despite different tissue contexts, we confirmed high directional concordance of the effect of chromatin accessibility on gene expression for peak-gene pairs significant in both datasets (mean Pearson *r* = 0.62 of effect sizes, 99% mean concordance across all the datasets: **Supplementary Figure 5b**). For comparison, we tested two popular linear parametric single-cell multimodal methods that are already published, namely ArchR^56^ or Signac^50^. Using arthritis-tissue dataset as a discovery and public PBMC as a replication, we noted lower directional concordance and effect correlation in these methods than in SCENT (mean Pearson’s *r* = 0.31, 62% mean directional concordance in ArchR and *r* = 0.24, 98% mean directional concordance in Signac; Supplementary Table 2b and **c**). These results argue that SCENT can more reproducibly detect enhancer-gene links compared with previous parametric methods for single-cell multimodal data.

To assess if SCENT peaks (i.e., *cis*-regulatory regions) were functional, we examined if (1) they co-localized with conventional *cis*-regulatory annotation, (2) their effect on expression was greater for closer peak-gene pairs, (3) they had high sequence conservation, and (4) peak-gene connections were more likely to be validated experimentally.

First, we tested the overlap of SCENT peaks with an ENCODE cCRE^63^, a conventional *cis*-regulatory annotation by bulk epigenomic datasets. We observed that 98.0% of the SCENT peaks overlapped with ENCODE cCRE on average, compared to 23.3% of random *cis*-regions matched for size and 89.0% of non-SCENT peaks (**Supplementary Figure 5c**).

Second, we examined the strength of enhance-gene links, hypothesizing that stronger links would be more proximal to the transcription start site (TSS) of target genes. The regression coefficient *β_peak_*(the effect size of peak accessibility on gene expression) became larger and more positive as the SCENT peaks got closer to the TSS (**Figure 2b** and **Figure 2c**, left panel), consistent with previous observations^56,64^. We annotated SCENT peaks with 18-state chromHMM results from 41 immune-related samples in ENCODE consortium^37^. When we subset peaks to those within enhancer annotations, we observed a clearer decay in *β_peak_* as a function of TSS distance (**Figure 2c**, right panel).

Third, we assessed whether SCENT peaks had higher sequence conversation across species, quantified as phastCons score^66^, which should indicate functional importance; the evolutionary conserved regulatory regions are known to be enriched for complex trait heritability^65^. As expected, exonic regions were much more evolutionary conserved than all non-coding *cis*-region (mean Δ phastCons score = 0.38, paired t-test *P* < 10^−323^; **Figure 2d**, purple). The SCENT regulatory regions were also conserved relative to non-coding *cis*-regions (mean Δ phastCons score = 0.13, paired t-test *P* = 4.2×10^−42^ in arthritis-tissue dataset; **Figure 2d**, green). In contrast, the Δ phastCons score between all *cis*-ATAC peaks and all non-coding *cis*-region was more modest (mean Δ phastCons score = 0.092, paired t-test *P* = 8.7×10^−27^ in arthritis-tissue dataset; **Figure 2d**, yellow). To test if the higher conservation in SCENT peaks were driven by their proximity to TSS (**Supplementary Figure 6a**), we matched each of the SCENT peak-gene pairs to one non-SCENT peak-gene pair that had the most similar TSS distance (**Supplementary Figure 6b**). We assessed Δ phastCons score between SCENT peaks and non-SCENT peaks with matching peaks on TSS distance. SCENT peaks had significantly higher conservation scores than the non-SCENT peaks with the matched TSS distance (mean Δ phastCons score = 0.034, *P* = 4.7×10^−4^ in arthritis-tissue dataset; **Supplementary Figure 5d**; see **Methods**). The higher sequence conservation suggested the functional importance of SCENT regulatory regions not solely driven by TSS proximity.

Finally, we tested whether the target genes from SCENT were enriched for experimentally confirmed enhancer-gene links. We used Nasser et al.^39^ CRISPR-Flow FISH results which included 278 positive enhancer-gene connections and 5,470 negative connections. The SCENT peaks were >4-fold enriched relative to non-SCENT peaks for positive connections (Fisher’s exact OR=4.5X, *P*=1.8×10^−9^ in arthritis-tissue dataset and 4.5X, *P*=1.0×10^−8^ in public PBMC dataset; **Methods,** Supplementary Table 3).

We anticipate that the genes with the largest number of SCENT peaks are likely to be the most constraint and least tolerant to loss of function mutations. The genes with the most SCENT peaks included *FOSB* (*n* = 97), *JUNB* (*n* = 95), and *RUNX1* (*n* = 77), critical and highly conserved transcription factors. We used mutational constraint metrics based on the absence of deleterious variants within human populations (i.e., the probability of being loss-of-function intolerant (pLI)^67^ and the loss-of-function observed/expected upper bound fraction (LOEUF)^68^). The normalized number of SCENT peaks per gene is strongly associated with mean constraint score for the gene (beta=0.37, *P*=4.9×10^−90^ for pLI where higher score indicates more constraint, and beta=-0.35, *P*=-0.35×10^−106^ for LOEUF where lower score indicates more constraint; **Supplementary Figure 7a** and **7b**, respectively). Previously, genes with many regulatory regions from bulk-epigenomic data had been shown to be enriched for loss-of-function intolerant genes^69^. We replicated the same trend in the single-cell multimodal datasets and SCENT.

### Enrichment of eQTL putative causal variants in SCENT peaks

We examined whether the SCENT peaks are likely to harbor statistically fine-mapped putative causal variants for expression quantitative loci (eQTL). We used tissue-specific eQTL fine-mapping results from GTEx across 49 tissues^70^ and defined any variants with posterior inclusion probability (PIP) > 0.2 as putative causal variants. We computed enrichment statistics within ATAC peaks or SCENT peaks (see **Methods**). Unsurprisingly, all the accessible regions defined by ATAC-seq in *cis*-regions were modestly enriched in fine-mapped variants by 2.7X (yellow, **Figure 3a**). However, SCENT peaks were more strikingly enriched in fine-mapped variants by 9.6X on average across all datasets (green, **Figure 3a**). Using more stringent PIP threshold cutoffs (0.5 and 0.7) to define putative causal variants resulted in even stronger enrichments (**Supplementary Figure 8**).

**Figure 3.**
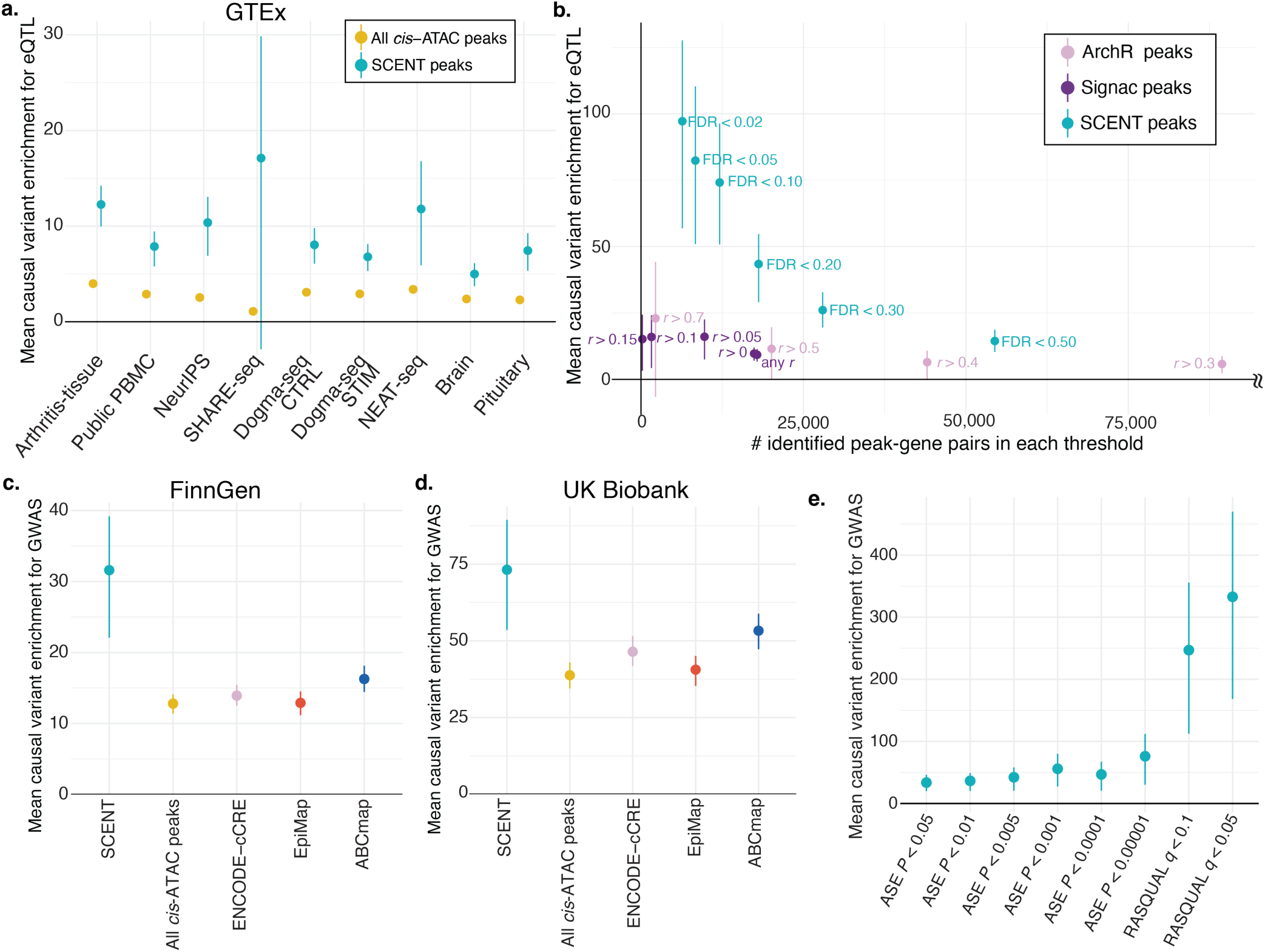
SCENT enhancers are enriched in putative causal variants of eQTL and GWAS. **a**. The mean causal variant enrichment for eQTL within SCENT peaks or all ATAC-seq peaks in each of the 9 single-cell datasets. The bars indicate 95% confidence intervals by bootstrapping genes. **b**. Comparison of the mean causal variant enrichment for eQTL (y-axis) between SCENT (green), ArchR (pink), and Signac (purple) as a function of the number of significant peak-gene pairs at each threshold of significance. The bars indicate 95% confidence intervals by bootstrapping genes. The ArchR results with > 100,000 peak-gene linkages are omitted, and full results are in Supplementary Figure 9b. **c** and **d**. The mean causal variant enrichment for GWAS within SCENT enhancers (green), all cis-ATAC peaks (yellow), ENCODE cCREs (pink), EpiMap enhancers across all groups (red) and ABC enhancers across all samples (blue). GWAS results were based on FinnGen (**c**) and UK Biobank (**d**). The bars indicate 95% confidence intervals by bootstrapping traits. **e**. The mean causal variant enrichment for FinnGen GWAS within intersection of SCENT enhancers and caQTL enhancers at each threshold of significance. The bars indicate 95% confidence intervals by bootstrapping traits.

Since many SCENT peaks are close to TSS regions, we again considered whether this enrichment might be driven by TSS proximity (**Supplementary Figure 6a**). To test this, we compared the fine-mapped variant enrichment between SCENT and non-SCENT peak-gene pairs with matched TSS distance (**Supplementary Figure 6b**). The SCENT peaks consistently had higher enrichment in all analyzed datasets (**Supplementary Figure 9a**) than TSS-distance-matched non-SCENT peaks (e.g., 12.3X in SCENT vs. 9.64X in distance-matched non-SCENT in arthritis-tissue dataset). This suggests that SCENT has additional information in identifying functional *cis*-regulatory regions beyond TSS distance.

We next compared the enrichment for eQTL putative causal variants in SCENT peaks to peaks identified by the two published linear parametric methods using single-cell multimodal data, ArchR^56^ and Signac^50^ using the same dataset. ArchR and Signac peaks had substantially lower causal variant enrichment for eQTL (1.4X and 9.3X, respectively) compared to SCENT peaks (74.1X) with FDR<0.10. We were concerned that this performance differences may reflect variable recall; that is SCENT may be more restrictive and calling fewer peaks. By varying the thresholds to define significant peak-gene associations (see **Methods**), we called the number of peak-gene pairs with difference levels of stringency and tested causal variant enrichment (i.e., recall-precision tradeoff; **Figure 3b** and **Supplementary Figure 9b**). SCENT peaks consistently demonstrated higher causal variant enrichment (i.e., precision) than ArchR and Signac peaks across different recall values.

We also tested Cicero^51^, which is a published linear parametric method for detecting promoter-enhancer co-accessibility from ATAC-seq data alone. We confirmed that SCENT peaks demonstrated higher causal variant enrichment than Cicero using the same dataset but only with ATAC-seq side (**Supplementary Figure 9c;** see **Methods**).

We assessed whether the Poisson regression or the bootstrapping in SCENT was driving its performance over other linear parametric methods. We benchmarked causal variant enrichment in SCENT peaks against peaks identified with only Poisson regression but without non-parametric bootstrapping (see **Methods**). As previously mentioned, we already observed false positive associations in the simulated null datasets in the Poisson-only strategy (**Supplementary Figure 1c**). Indeed, we observed substantially lower causal variant enrichment at a given recall compared to SCENT (14.4X in Poisson only vs. 74.1X in SCENT at the same FDR<0.10), albeit slightly higher than the linear methods ArchR and Signac (**Supplementary Figure 9c**). This underscored the importance of accounting for both (1) sparsity by Poisson regression and (2) highly variable gene count distribution by non-parametric bootstrapping to achieve high precision in SCENT.

SCENT can detect *cis*-regulatory regions in a cell-type-specific manner. We created cell-type-specific enhancer-gene maps in four major cell types with > 5,000 cells across datasets; for each cell type we took the union of SCENT enhancers across datasets. The cell-type-specific SCENT enhancers (e.g., SCENT B cell peaks) were most enriched in putative causal eQTL variants within relevant samples in GTEx (e.g., EBV-transformed lymphocytes; **Supplementary Figure 9d**).

These results suggest that SCENT can prioritize regulatory elements harboring putative causal eQTL variants in a cell-type-specific manner, with higher precision than the previous single-cell methods.

### Enrichment of likely causal variants for GWAS in SCENT enhancers

SCENT applied for multimodal data from disease-relevant tissues can build disease-specific enhancer-gene maps. We sought to examine whether SCENT peaks can be used for the more difficult task of prioritizing disease causal variants. We obtained candidate causal variants for diseases and traits from fine-mapping results of GWASs in two large-scale biobanks (PIP>0.2; FinnGen^71^ [1,046 disease traits] and UK Biobank^72^ [35 binary traits and 59 quantitative traits])^28^. We computed enrichment statistics for causal GWAS variants within SCENT enhancers (both cell-type-specific tracks and aggregated tracks across cell types; see **Methods**). The SCENT enhancers were strikingly enriched in causal GWAS variants in FinnGen (31.6X on average; 1046 traits; **Figure 3c** and **Supplementary Figure 10a**) and UK Biobank (73.2X on average; 94 traits; **Figure 3d** and **Supplementary Figure 10b**). This enrichment was again much larger than all *cis*-ATAC peaks (12.8X in FinnGen and 38.8X in UK Biobank). Moreover, the target genes of the likely causal variants for autoimmune diseases (AID) identified by SCENT peaks in immune-related cell types had higher fraction (10.8%) of know genes implicated in Mendelian disorders of immune dysregulation (*n*_gene_ = 550)^73,74^ than SCENT peaks in fibroblast (3.8%; **Supplementary Figure 10c**).

We compared SCENT to alternative genome annotations and enhancer-gene maps from bulk tissues. Causal variant enrichment in SCENT was much higher than the conventional bulk-based annotations such as ENCODE cCREs (13.9X in FinnGen and 46.5X in UK Biobank), ABC (16.3X in FinnGen and 53.3X in UK Biobank) and EpiMap (12.9X in FinnGen and 40.6X in UK Biobank; **Figure 3c** and **3d** [aggregated tracks], **Supplementary Figure 10a** and **10b** [cell-type-specific tracks]). We again assessed recall and precision tradeoffs by varying thresholds for defining significant peak-gene linkages. We constructed SCENT from 9 datasets and 23 cell types with only 28 samples, substantially less than the 833 samples and tissues used to construct EpiMap and 131 samples and cell lines for the ABC model. Despite the smaller data set, SCENT peaks consistently demonstrated higher precision (i.e., enrichment of causal GWAS variants) at a given recall (i.e., a similar number of identified peak-gene linkages) than ABC model and EpiMap (**Supplementary Figure 11a**). A more stringent PIP threshold (0.5 and 0.7) for putative causal variants increased the enrichment while maintaining the higher enrichment in SCENT than bulk methods (**Supplementary Figure 11b**). The target genes for AID by SCENT in immune-related cell types had higher fraction (10.8%) of known Mendelian genes of immune dysregulation^73,74^ than EpiMap (8.6%) and ABC model (4.4%) (**Supplementary Figure 10c**). These results demonstrate the power SCENT achieved by accurately modeling association between chromatin accessibility and gene expression at the single-cell resolution.

We hypothesized that putative causal variants by SCENT would likely modulate chromatin accessibility (e.g., transcription factor binding affinity). If so, the intersection of the SCENT enhancers and chromatin accessibility quantitative trait loci (caQTL) could further enrich the causal GWAS variants^75–78^, because these intersected enhancers should include genetic variants that directly change both chromatin accessibility and gene expression. To test this hypothesis, we used single-cell ATAC-seq samples with genotype (*n*_donor_ = 17; arthritis-tissue dataset) and performed caQTL mapping by leveraging allele-specific (AS) chromatin accessibility (binomial test followed by meta-analysis across donors) or by combining AS with inter-individual differences (RASQUAL^79^). We then intersected the caQTL ATAC peaks with the SCENT enhancers and calculated the causal variant enrichment within these intersected regions. We observed higher enrichment within intersected regions with SCENT and caQTL than those with SCENT alone. The enrichment increased as we used more stringent threshold for caQTL peaks, reaching as high as 333-fold when compared with background *cis*-regions (**Figure 3e**). Thus, SCENT efficiently prioritized causal GWAS variants in part by capturing regulatory regions of which chromatin accessibility is perturbed by genetic variants and modulates gene expression. SCENT demonstrated a potential to further enrich causal variants by caQTLs if multimodal data has matched genotype data.

### Defining mechanisms of GWAS loci by SCENT

We finally sought to use SCENT enhancer-gene links to define disease causal mechanisms. We analyzed the fine-mapped variants from GWASs (FinnGen, UK Biobank and GWAS cohorts of rheumatoid arthritis (RA)^26^, inflammatory bowel disease^29^ and type 1 diabetes (T1D)^80^). SCENT linked 4,124 putative causal variants (PIP>0.1) to their potential target genes across 1,143 traits (Supplementary Table 4). These target genes were mostly close to the causal variant, with 20% of them being the closest gene to the causal variant (**Supplementary Figure 12a** and **12b**; see **Methods**). However, 30.6% of the time SCENT linked causal variants to genes more than 300 kb away.

We first focus on autoimmune loci, given that our current SCENT tracks are largely derived from immune cell types. We prioritized a single well fine-mapped variant rs72928038 (PIP > 0.3) at 6q15 locus in multiple autoimmune diseases (RA, T1D, atopic dermatitis and hypothyroidism), within the T-cell-specific SCENT enhancer (T cells in Public PBMC and Dogma-seq datasets; **Figure 4a**). This enhancer was linked to *BACH2*, which was also the closest gene to this fine-mapped variant. Notably, base-editing in T cells has confirmed that this variant affects *BACH2* expression^81^. Moreover, editing of this variant into CD8 T cells skewed naive T cells toward effector T cell fates^81^.

**Figure 4.**
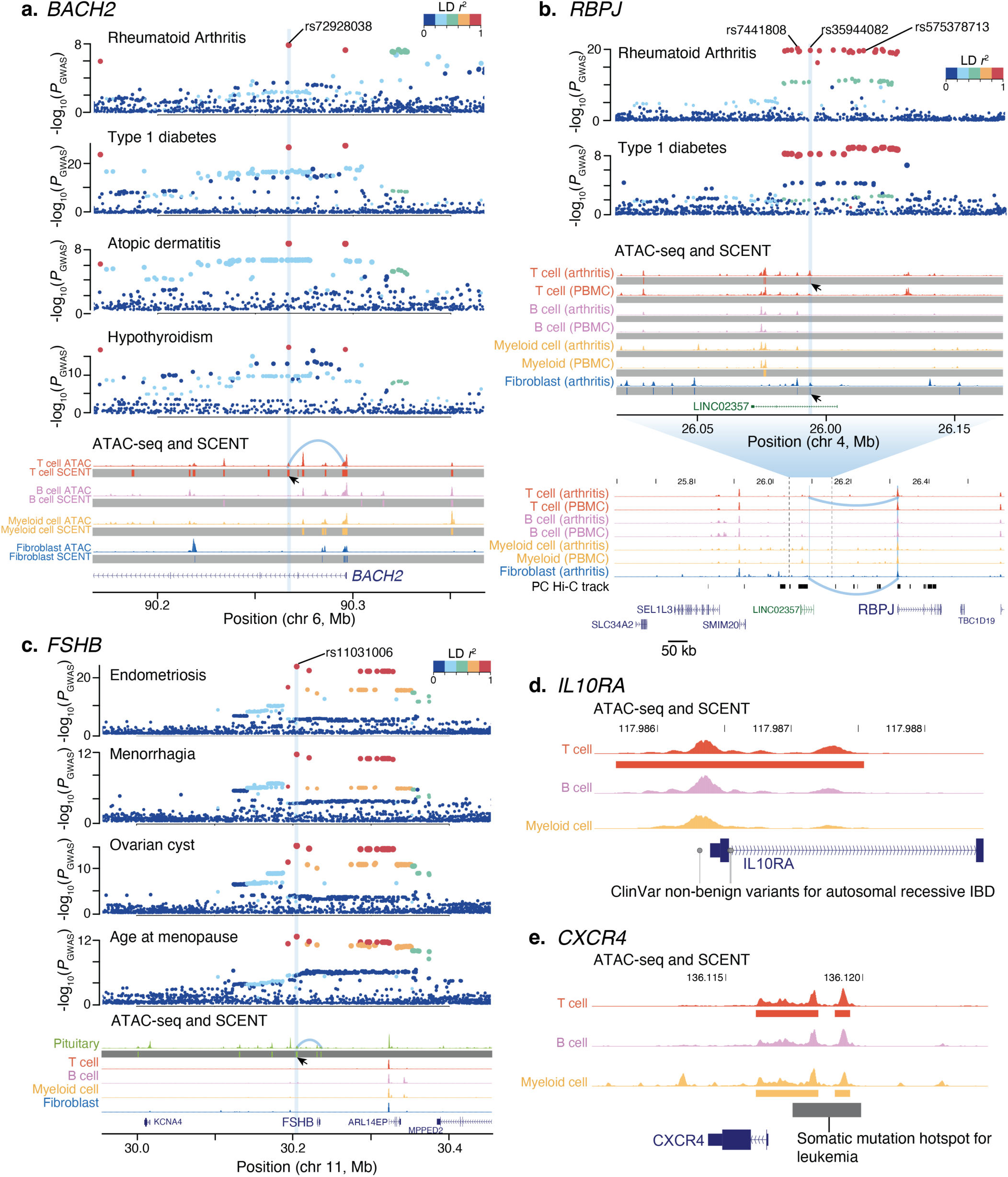
SCENT defined causal variants and genes in complex trait GWAS. **a**. Rs72928038 at *BACH2* locus was prioritized by T-cell-specific SCENT enhancer-gene map, being for RA, T1D, Atopic dermatitis and hypothyroidism. The top four panels are GWAS regional plots, with x-axis representing the position of each genetic variant. The color of the dots represent LD *r*^2^ from the prioritized variant (highlighted by light blue stripe). ATAC-seq and SCENT tracks represent aggregated ATAC-seq tracks (top) and SCENT peaks (bottom with grey stripes) in each cell type (public PBMC dataset for immune cell types and arthritis-tissue dataset for fibroblast). An arrow head indicates the SCENT peak overlapping with fine-mapped variant. **b**. Rs35944082 for RA and T1D was prioritized and connected to *RBPJ* by long-range interaction from T-cell-and fibroblast-SCENT enhancer-gene map using inflamed synovium in arthritis-tissue dataset. The top two panels are GWAS regional plots similarly to panel **a**. ATAC-seq and SCENT tracks are shown similarly to panel **a**, but using both public PBMC and arthritis-tissue datasets. **c**. Rs11031006 was prioritized and connected to *FSHB* for multiple gynecological traits by using pituitary-derived single-cell multimodal dataset. The top four panels are GWAS regional plots similarly to panel **a**. ATAC-seq and SCENT tracks are shown similarly to panel **a**, and include tracks from pituitary dataset. There were no SCENT peaks in cell types except for pituitary. **d**. ATAC-seq and SCENT tracks for *IL10RA* locus, where non-coding ClinVar variants (grey dots) colocalized with T-cell SCENT track. **e**. ATAC-seq and SCENT tracks for *CXCR4* locus, where somatic mutation hotspot for leukemia colocalized with T-cell and myeloid-cell SCENT tracks.

Another locus for RA and T1D at 4p15.2 harbored 21 candidate variants, each with low PIPs (< 0.14). SCENT prioritized a single variant rs35944082 in T cells and fibroblasts only within the arthritis-tissue dataset from inflamed synovial tissue (**Figure 4b**). SCENT linked this variant to *RBPJ*, which was the 3rd closest gene to this variant located 235kb away. This variant-gene link was supported by a physical contact from promotor-capture Hi-C data in hematopoietic cells^82^. *RBPJ* (recombination signal binding protein for immunoglobulin kappa J region) is a transcription factor critical for NOTCH signaling, which has been implicated in RA tissue inflammation through functional studies^83,84^. *Rbpj* knockdown in mice resulted in abnormal T cell differentiation and disrupted regulatory T cell phenotype^85,86^, consistent with a plausible role in autoimmune diseases. Intriguingly, we observed no SCENT peaks in T cells from PBMC or blood at this locus. This linkage was not present in EpiMap. ABC map prioritized another variant, rs7441808 at this locus and linked it non-specifically to 16 genes including *RBPJ*, making it difficult to define the true causal gene. These results underscored the importance of creating enhancer-gene links using causal cell types, in this case cells from inflammatory tissues, in the instances where links exist only in disease-relevant tissues.

We highlight another example of SCENT to build enhancer-gene maps from disease-critical tissues. We examined the enhancer-gene map produced from single-cell pituitary data^62^ to assess 11p14.1 locus for multiple gynecological traits (endometriosis, menorrhagia, ovarian cyst and age at menopause). Our map connected rs11031006 to *FSHB* (follicle stimulating hormone subunit beta) (**Figure 4c**), which is specifically expressed in the pituitary^70,87^ and enables ovarian folliculogenesis to the antral follicle stage^88^. Rare genetic variants within *FSHB* cause autosomal recessive hypogonadotropic hypogonadism^89^. However, multimodal data from other tissues and bulk-based methods (ABC model and EpiMap) were unable to prioritize this variant, since they missed the most disease-relevant tissue of pituitary.

### Mendelian-disease variants and somatic mutations in cancer within SCENT enhancers

Having established the SCENT’s utility in defining likely causal variants and genes in complex diseases, we examined rare non-coding variants causing Mendelian diseases. Currently, causal mutations and genes can only be identified in ~30–40% of patients with Mendelian diseases^90–92^. Consequently, many variants in cases are annotated as variants of uncertain significance (VUS). The VUS annotation is especially challenging for non-coding variants. We examined the overlap of clinically reported non-benign non-coding variants by ClinVar^93^ (400,300 variants in total) within SCENT enhancers. The SCENT enhancers harbored 2.0 times ClinVar variants on average than all the ATAC regions with the same genomic length across all the datasets (**Supplementary Figure 13**). This density of ClinVar variants was 3.2 times and 12 times on average larger than that in ENCODE cCREs and of all non-coding regions, respectively. We defined 3,724 target genes for 33,618 non-coding ClinVar variants by SCENT in total (Supplementary Table 5). As illustrative examples, we found 40 non-coding variants linked to *LDLR* gene causing familial hypercholesterolemia 1^93^, 3 non-coding variants linked to *IL10RA* causing autosomal recessive early-onset inflammatory bowel disease 28 (**Figure 4d**)^94^, and an intronic variant rs1591491477 linked to *ATM* gene causing hereditary cancer-predisposing syndrome^93^.

Finally, we used SCENT to connect non-coding somatic mutation hotspots to target genes. Recently, somatic mutation analyses across the entire cancer genome revealed possible driver non-coding events^95^. Among 372 non-coding mutation hotspots in 19 cancer types, SCENT enhancers included 193 cancer-mutation hotspot pairs (Supplementary Table 6). SCENT enhancer-gene linkage successfully linked those hotspots to known driver genes (e.g., *BACH2*, *BCL6*, *BCR*, *CXCR4* (**Figure 4e**), and *IRF8* in leukemia). In some instances, SCENT nominated different target genes for these mutation hotspots from those based on ABC model used in the original study. For example, SCENT connected a somatic mutation hotspot in leukemia at chr14:105568663-106851785 to *IGHA1* (Immunoglobulin Heavy Constant Alpha 1), which might be more biologically relevant than *ADAM6* nominated by ABC model. These results implicate broad applicability of SCENT for annotating all types of human variations in non-coding regions.

### Augmenting SCENT enhancer-gene maps with more samples

While the recall for enhancer-gene maps defined by SCENT was lower than that by bulk-tissue-based methods, this might be a function of current limited sample sizes. We assessed if the addition of more cells into SCENT leads to the higher recall for enhancer-gene maps while retaining the precision. By downsampling of our multimodal single cell dataset, we observed that the number of significant gene-peak pairs increased linearly to the number of cells per cell type in a given dataset, suggesting that SCENT will be even better powered as the size of sc-multimodal datasets increases (**Supplementary Figure 14**). We considered the possibility that enhancer-gene maps with greater numbers of cells might capture spurious associations; if this was the case, we would expect more long-range associations, which are more likely to be false positives with greater cell numbers. In contrast, shorter-range and longer-range associations were both equivalently represented as we added cells, suggesting the robustness of our discovery.

## Discussion

In this study, we presented a novel statistical method, SCENT, to create a cell-type-specific enhancer-gene map from single-cell multimodal data. Single-cell RNA-seq and ATAC-seq are both sparse and have variable count distributions, which requires non-parametric bootstrapping to connect chromatin accessibility with gene expression. The SCENT model demonstrated well-controlled type I error, outperforming commonly used statistical models which showed inflated statistics. SCENT mapped enhancers that showed strikingly high enrichment for putative causal variants in eQTLs and GWASs and outperformed previous methods for single-cell multimodal data (e.g., ArchR^49^ and Signac^50^). Despite using substantially lower number of samples (28 from 9 datasets in total), enhancers defined by SCENT had equivalent or even higher enrichment for putative causal variants than bulk-tissue-based methods with more than 100 samples (e.g., EpiMap and ABC model), by modeling single-cell level observations instead of obscuring them into sample-level association.

As potential limitations, first, our enhancer-gene maps had relatively fewer enhancers (lower recall) compared to other resources (**Figure 2a**). However, downsampling experiments showed a clear linear relationship between the number of cells and the number of significant SCENT peak-gene links. It follows that SCENT applied to larger datasets from a diverse set of tissues will further expand the current enhancer-gene map. In contrast, bulk-tissue-based enhancer-gene map might have an upper limit of discovery by the number of samples generated by each consortium (e.g., ENCODE). Second, SCENT focuses on gene *cis*-regulatory mechanisms to fine-map disease causal alleles, while there could be other causal mechanisms that explain disease heritability, such as alleles that act through *trans*-regulatory effects, splicing effects, or post-transcriptional effects^96^.

We argue that the real utility of SCENT is that it enables the construction of disease-tissue-relevant enhancer-gene maps. Multimodal single cell data can be easily obtained from a wide range of primary human tissues. Since these assays query nuclear material, data can be obtained without disaggregating tissues and thus can be employed for assays that need intact cells from tissue. Therefore, it is possible to build relevant tissue-specific enhancer-gene maps that are necessary to understand the causal mechanisms of common diseases, rare diseases, and somatic non-coding mutations in cancers. For example, understanding the *FSHB* locus in gynecological traits specifically required a pituitary map, and *RBPJ* locus in RA specifically required a synovial tissue map.

In summary, our method SCENT is a robust, versatile method to efficiently define causal variants and genes in human diseases and will fill the gap in the current enhancer-gene map built from genomic data in bulk tissues.

## Data Availability

The publicly available datasets were downloaded via Gene Expression Ombibus (accession codes: GSE140203, GSE156478, GSE178707, GSE193240, GSE178453) or web repository (https://www.10xgenomics.com/resources/datasets?query=&page=1&configure%5Bfacets%5D%5B0%5D=chemistryVersionAndThroughput&configure%5Bfacets%5D%5B1%5D=pipeline.version&configure%5BhitsPerPage%5D=500&menu%5Bproducts.name%5D=Single%20Cell%20Multiome%20ATAC%20%2B%20Gene%20Expression, https://openproblems.bio/neurips_docs/data/dataset/). The raw data for arthritis-tissue dataset (single-cell multimodal RNA/ATAC-seq and single-cell ATAC-seq) will be publicly available before the acceptance of this manuscript.

## Code Availability

The computational scripts related to this manuscript are available at https://github.com/immunogenomics/SCENT.

## Methods

### Data and sample in arthritis-tissue dataset

This study was performed in accordance with protocols approved by the Brigham and Women’s Hospital and the Hospital for Special Surgery institutional review boards. Synovial tissue from patients with RA and OA were collected from synovectomy or arthroplasty procedures followed by cryopreservation as previously described^97^. RA samples with high levels of lymphocyte infiltration (as scored by a pathologist on histologic sections) were identified as “inflamed” and used for downstream analysis. Next, cryopreserved synovial tissue fragments were dissociated by a mechanical and enzymatic digestion^97^, followed by flow sorting to enrich for live synovial cells. For each tissue sample, the viable cells were isolated and lysed to extract and load approximately 10,000 nuclei according to manufacturer protocol (10X Genomics). Joint sc-RNA- and sc-ATAC-seq libraries were prepared using the 10x Genomics Single Cell Multiome ATAC + Gene Expression kit according to manufacturer’s instructions. Libraries were sequenced with paired-end 150-bp reads on an Illumina Novaseq to a target depth of 30,000 read pairs per nuclei both for mRNA and ATAC libraries. Demultiplexed scRNA-seq fastq files were inputted into the Cell Ranger ARC pipeline (version 2.0.0) from 10x Genomics to generate barcoded count matrix of gene expression. For ATAC-seq, we trimmed adaptor and primer sequences and mapped the trimmed reads to the hg38 genome by BWA-MEM with default parameters. To deduplicate reads from PCR amplification bias within a cell while keeping reads originating from the same positions but from different cells, we used in-house scripts (manuscript in preparation).

### Uniform processing of single-cell multimodal datasets

In addition to our arthritis-tissue multimodal dataset, we downloaded all publicly available multimodal RNA-seq/ATAC-seq datasets from adult human tissues (*n*_dataset_ = 9, as of April 2022). We processed these downloaded count matrices of gene expression and ATAC data. Briefly, we applied QC to both the nuclear RNA data and the ATAC data based on RNA counts, ATAC fragments, nucleosome signal, and TSS enrichment (Supplementary Table 7). We only kept cells that had passed QC in both RNA-seq and ATAC-seq. Then to identify open chromatin regions (peaks), we used macs2 to call open chromatin peaks using post-QC ATAC-seq data. We thus obtained count matrices of gene expression and ATAC peaks with corresponding cell barcodes. Gene expression counts were normalized using the NormalizeData function (Seurat^98^), scaled using the ScaleData function (Seurat), and batch corrected using Harmony^99^. We visualized the cells in two low-dimensional embeddings with UMAP by using 20 batch-corrected principal components from these normalized gene expression matrices (**Figure 1c**). When original cell labels are provided by the authors, we used those labels to obtain broad cell type categories. When they are not available, we performed reference-query mapping by Seurat and PBMC reference object to define broad cell type labels. ATAC peak matrix was binarized to have 1 if a count is > 0 and 0 otherwise.

### SCENT method

We defined *cis*-peaks as any peaks whose center is within the window +/-500 kb from a given gene body. We modeled the association between peak’s binarized accessibility and the target gene’s expression with Poisson distribution:

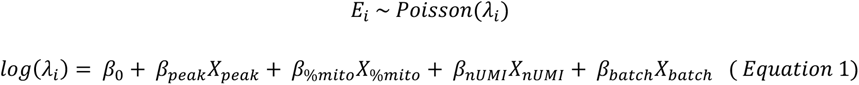

where *E_i_* is the observed expression count of *i*th gene, and *λ_i_* is the expected count under Poisson distribution. *β_peak_* indicates the effect of chromatin accessibility of a peak on *i* th gene expression. *Β_%mito_*, *β_nUMI_*, and *β_batch_* each represents the effect of covariates, percentage of mitochondrial reads per cell as a mesure of cell quality, the number of UMIs in the cell, and the batch, respectively. To empirically assess error and significance of *β_peak_* for each peak-gene combination, we used bootstrapping procedures. In brief, we resampled cells with replacement in each bootstrapping procedure and re-estimated *β′_peak_* within those resampled cells. We repeated this procedure *N* times, where we adaptively increased *N* (i.e., the total number of bootstrapping) from at least 100 and up to 50,000, depending on the significance of *β_peak_* (as described next) in each chunk of bootstrapping trials to reduce the computational burden. After *N* times of bootstrapping, we assessed the distribution of *N β′_peak_*s against null hypothesis (*β′_peak_* = 0) to derive the significance of *β_peak_* (i.e., two-sided bootstrapping-based *P* value for this peak-gene combination by counting the instances where the statistics are equal or more extreme than the null hypothesis of *β′_peak_* = 0; **Supplementary Figure 2**).

To avoid spurious associations from rare ATAC peak and rare gene expression, we QCed cis-peak-gene pairs we test so that both peak and gene should have been expressed in at least 5% of the cells we analyze. We finally defined a set of significant peak-gene pairs for each cell type based on bootstrapping-based *P* values and FDR correction for multiple testing (Benjamini & Hochberg correction).

When we tested the calibration of statistics from SCENT or other regression strategies (**Supplementary Figure 1**), we used null dataset where we randomly permuted cell labels in the ATAC-seq and ran the regression model we tested.

### ArchR peak2gene and Signac LinkPeaks method

We analyzed arthritis-tissue dataset with ArchR^49^ and Signac^50^ for single-cell multimodal data, which both have a function to define peak-gene linkages. In brief, ArchR takes multimodal data and creates low-overlapping aggregates of single cells based on *k*-nearest neighbor graph. Then it correlates peak accessibility with gene expression by Pearson correlation of aggregated and log2-normalized peak count and gene count. Signac computes the Pearson correlation coefficient *r* (corSparse function in R) for each gene and for each peak within 500kb of the gene TSS. Signac then compares the observed correlation coefficient with an expected correlation coefficient for each peak given the GC content, accessibility, and length of the peak. Signac defines *P* value for each gene-peak links from the z score based on this comparison. We ran both methods on arthritis-tissue dataset with default parameters. We output statistics for all peak-gene pairs we tested without any cut-off for correlation *r* or *P* values. We used FDR in the output from ArchR software, or computed FDR using *P* values in the output from Signac software by Benjamini & Hochberg correction. We defined significant peak-gene linkages as those with FDR < 0.10, and used varying correlation *r* to assess the precision and recall in the causal variant enrichment analysis (see later sections in **Method**).

### Replication across datasets

Since we have the same immune-related cell types across different multimodal datasets, we evaluated the concordance of enhancer-gene map in a discovery dataset (arthritis-tissue dataset) when compared with other replication datasets including immune-related cell types (Public PBMC, NeurIPS, SHARE-seq and NEAT-seq datasets). To this end, we used most stringent FDR threshold for defining an enhancer-gene map in arthritis-tissue dataset (FDR < 1%). We then used more lenient threshold for defining an enhancer-gene map in replication datasets (FDR < 10%), which is a similar strategy used in assessing replication in GWAS. For each cell type and for each replication dataset, we took the intersection of enhancer-gene links defined as significant in both datasets. We assessed the directional concordance (i.e., concordance of the sign of *β_peak_*) and the Pearson’s correlation *r* of *β_peak_* between the discovery and the replication for these peak-gene pairs. For the largest replication dataset of Public PBMC, we performed the same analysis for enhancer-gene map from ArchR and Signac software.

### Conservation score analysis

To compare the evolutional conservation across species between our annotated peaks and the other peaks, we used phastCons^66^ score. We downloaded the phastCons score for multiple alignments of 99 vertebrate genomes from https://hgdownload.cse.ucsc.edu/goldenpath/hg19/phastCons100way/. We lifted them over to GRCh38 by LiftOver software. We used SCENT results for arthritis-tissue, Public PBMC and NeurIPS for conservation score analysis as representative datasets with the largest numbers of cells. Because each gene should have variable functional importance and conservation, we assessed each gene separately. For each gene, we took (1) an annotation of interest for the gene and (2) all *cis*-non-coding regions (< 500kb from a gene), and computed the mean phastCons score of each of two sets of the peaks. As annotations to be tested, we used a. exonic regions of the gene, b. SCENT peaks for the gene, and c. all ATAC peaks in cis-regions from the gene (< 500 kb). Then, we took the difference between two mean differences (Δ phastCons score), and computed the mean differences across all the genes (mean Δ phastCons score) as follows.

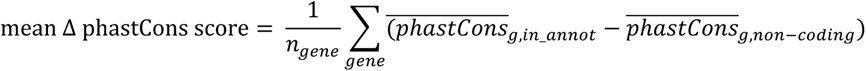

By bootstrapping the genes, we calculated the 95% CI of the mean Δ phastCons score. If this metric is positive, that indicates that the annotated regions are more conserved than non-coding regions.

We also calculated similar Δ phastCons score by comparing the SCENT peaks with TSS-distance-matched non-SCENT peaks in each dataset.

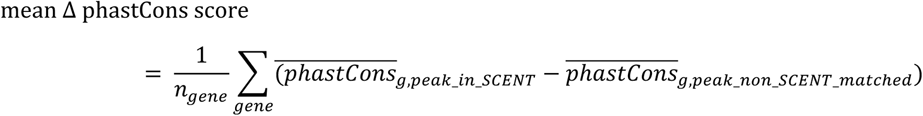

By bootstrapping the genes, we again calculated the 95% CI of the mean Δ phastCons score. If this metric is positive, that indicates that SCENT peaks are more conserved than TSS-distance-matched non-SCENT peaks.

### Construction of a set of TSS-matched non-SCENT peaks

To assess the effect of TSS distance when comparing SCENT peaks with non-SCENT peaks, we matched each one of the SCENT peak-gene pairs to one non-SCENT peak-gene pair, where the peak had the most similar TSS distance to the same gene among all the ATAC peaks in *cis* in each of the dataset. We confirmed that the resulting TSS-distance-matched non-SCENT peak-gene pairs demonstrated the similar distributions of TSS distance when compared with the SCENT peak-gene pairs (**Supplementary Figure 6b**).

### Gene’s constraint and the number of significant SCENT peaks for a gene

We sought to investigate the relationship between the number of significant SCENT peaks for each gene and the gene’s evolutionary constraint. We used pLI and LOEUF as metrics for the gene’s loss-of-function intolerance within human population. We downloaded both pLI and LOEUF scores from gnomAD browser (https://gnomad.broadinstitute.org/downloads). We inverse-normal transformed the raw number of significant SCENT peaks for each gene, since the raw number of significant SCENT peaks for each gene is skewed toward zero (**Supplementary Figure 5a**). We performed linear regression between the normalized number of significant SCENT peaks and pLI or LOEUF score with accounting for gene length, which could be potential confounding factor for pLI and LOEUF^67,68^.

### Validation with CRISPR-Flow FISH results

To validate our SCENT enhancer-gene links, we used published CRISPR-Flow FISH experiments as potential ground-truth positive enhancer element-gene links and negative enhancer element-gene links. We downloaded the experimental results from the Supplementary Table 5 of original publication^39^. We used “Perturbation Target” as candidate enhancer elements. We defined 283 positive enhancer element-gene links when they are “TRUE” for “Regulated” column (i.e., the element-gene pair is significant and the effect size is negative) and 5,472 negative enhancer element-gene links when they are “FALSE” for “Regulated” column. We lifted them over to GRCh38 and obtained final sets of 278 positive links and 5,470 negative links.

We used two most powered datasets, arthritis-tissue and Public PBMC datasets. For each dataset, we used “bedtools intersect” to categorize SCENT peak-gene links and non-SCENT ATAC peak-gene pairs into either CRISPR-positive or CRISPR-negative groups, based on whether these peaks overlapped with positive or negative CRISPR-Flow FISH links for the same gene (Supplementary Table 3). We finally performed two-sided Fisher’s exact test to assess the enrichment of CRISPR-positive links within SCENT peak-gene links in each dataset.

### Cell-type-specific SCENT tracks and aggregated SCENT tracks

For cell types with more than 5,000 cells across datasets, we concatenated SCENT peak-gene linkages across all the datasets to create cell-type-specific SCENT tracks. We collected a set of SCENT peak-gene linkages for the same cell type and used “bedtools merge” function (for each gene) to obtain a union of SCENT peaks for each gene. Similarly, we created aggregated SCENT tracks across all the cell types and all datasets. We collected all sets of SCENT peak-gene linkages and used “bedtools merge” function (for each gene) to obtain a union of SCENT peaks for each gene across all the cell types and all datasets.

### Causal variant enrichment analysis using eQTLs

We defined a causal enrichment for eQTL within SCENT enhancers and other annotations by using statistically fine-mapped variant-gene combinations from GTEx. We used publicly available statistics analyzed by CAVIAR software^20^, and selected variants with PIP > 0.2 as putatively causal (fine-mapped) variants for primary analyses. For the primary enrichment analysis, we aggregated fine-mapped variants from all the 49 tissues. For cell-type-specific SCENT enrichment analysis (**Supplementary Figure 9d**), we used fine-mapped variants from each tissue separately. We intersected these putatively causal variants with our annotation (SCENT peaks, ArchR peaks or Signac peaks). We then retained any variants which the linking method (SCENT, ArchR, Signac, and Cicero) connected to the same gene as GTEx phenotype gene.

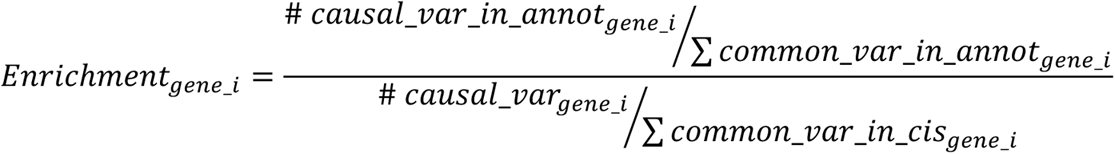

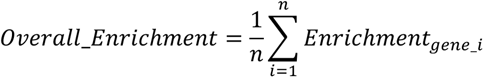

For each gene *i* (expression phenotype), we divided the number of putatively causal variants within an annotation normalized by the number of common variants within an annotation by the number of all causal variants for gene *i* normalized by the number of all common variants within cis-region from for gene *i*. To calculate common variants within annotation or within locus, we used 1000 Genomes Project genotype. We selected any variants with minor allele frequency > 1% in European population as a set of common variants to be intersected with each annotation. To derive *Overall_Enrichment* score, we took the mean across all the genes.

To have further insights into precision and recall and compare against ArchR peak2gene and Signac LinkPeaks functions, we varied the threshold for defining a set of significant peak-gene linkages in each software (i.e., FDR in SCENT {0.50, 0.30, 0.20, 0.10, 0.05, 0.02}, Peason’s correlation *r* {any, 0, 0.1, 0.3, 0.5, 0.7} in ArchR, and correlation socre {any, 0, 0.05, 0.1, 0.15} in Signac). We used the same myeloid cells in the arthritis-tissue dataset and a set of eQTL fine-mapped variants in GTEx blood tissue for this benchmark across all three methods. We then used each set of peak-gene linkages to re-calculate causal variant enrichment *Overall_Enrichment* score (**Figure 3b**).

We also assessed the impact of PIP threshold in defining a set of statistically fine-mapped variants on the causal variant enrichment analysis. To do so, we re-defined the set of putative causal variants with more stringent PIP thresholds (PIP > 0.5 and PIP > 0.7), and re-computed the calculate causal variant enrichment *Overall_Enrichment* score.

### Cicero co-accessibility analyses

To benchmark our SCENT using single-cell multimodal ATAC/RNA-seq against a published method using single-cell unimodal ATAC-seq alone, we ran Cicero^51^ for the same dataset of myeloid cells in the arthritis-tissue dataset as benchmarked in the SCENT, ArchR and Signac. We only used the peak by cell matrix from the ATAC-seq side of the arthritis-tissue dataset and ran “run_cicero” function with default parameters to obtain Cicero co-accessibility scores. We only retained peak-peak co-accessibility as potential enhancer-gene connection when one of the co-accessible peaks is a promoter of a gene (defined by the peak’s distance to the TSS < 1kb); we treated them as putative enhancer-gene (promoter) linkage. We used the co-accessibility scores {any, 0, 0.1, 0.3, 0.4, 0.5, 0.7} for assessing the recall-precision tradeoffs as described in the previous section.

### Peak-gene linkage using Poisson regression alone

As other benchmarking for assessing the effect of the components of SCENT on the causal variant enrichment, we also created peak-gene linkage using the Poisson regression but without non-parametric bootstrapping for the same dataset of myeloid cells in the arthritis-tissue dataset. We used the nominal *P* values for the term *X_peak_* from the Poisson regression (Equation (1)) to perform FDR correction to obtain significant peak-gene pairs using the Poisson regression alone. We then used the FDR thresholds {0.30, 0.20, 0.10, 0.05, 0.02, 0.01} for assessing the recall-precision tradeoffs as described in the previous section.

### GWAS fine-mapping results

We used GWAS fine-mapping results in FinnGen release 6^71^ upon registration and publicly available GWAS fine-mapping results in UK Biobank^72^ (https://www.finucanelab.org/data). For FinnGen traits, we downloaded all the fine-mapping results by SuSIE software^22^ and systematically selected any traits with case count > 1,000. We then selected non-coding fine-mapped loci which did not include any non-synonymous or splicing variants with PIP > 0.5. We thus analyzed 1,046 traits and 5,753 loci in total after QC. For UK Biobank, we analyzed the fine-mapping results by SuSIE software for all 94 traits including binary and quantitative traits. Since the genomic coordinates for the UK Biobank fine-mapping results were hg19, we lifted them over to GRCh38 by using LiftOVer software. We again selected non-coding fine-mapped loci which did not include any non-synonymous or splicing variants with PIP > 0.5. We thus analyzed 7,274 loci in total after QC.

We analyzed three additional autoimmune GWAS fine-mapping results for RA^26^, T1D^80^, and IBD^29^, given our special interest in immune-mediated traits. We similarly selected non-coding fine-mapped loci which did not include any non-synonymous or splicing variants with PIP > 0.5, and lifted the results over to GRCh38 by using LiftOVer software. We defined 117 loci for RA, 77 loci for T1D and 86 loci for IBD.

### Causal variant enrichment analysis using GWASs

We defined a causal enrichment for GWAS within SCENT enhancers and other annotations by using statistically fine-mapped variants from FinnGen^71^ and UK Biobank^72^ which we described in the previous section. We selected variants with PIP > 0.2 as putatively causal variants for primary analyses.

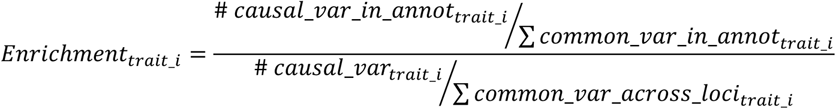

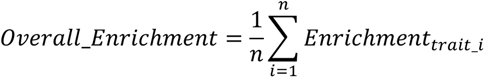

For each trait *i*, we divided the number of putatively causal variants within an annotation (across all loci for trait *i*) normalized by the number of common variants within an annotation by the number of all causal variants for trait *i* normalized by the number of all common variants within all significant loci analyzed for the trait *i*. To calculate common variants within annotation or within locus, we again used 1000 Genomes Project variants with minor allele frequency > 1% in European population. To derive *Overall_Enrichment* score, we took the mean across all the traits.

For each trait *i* and putative causal gene pair, we calculated the distance between the TSS of the gene and the most likely causal variant which had the largest PIP when multiple variants were nominated for a single gene by SCENT (**Supplementary Figure 12a**). For each putative causal gene for the trait *i*, we also sorted all the genes based on the distance between the gene’s TSS and the most likely causal variant (from the smallest to the largest). We then obtained the rank of the putative causal gene from SCENT among the sorted gene list to see how often the SCENT gene is the closest gene from the most likely causal variant.

### Comparison with bulk-tissue-based regulatory annotation and enhancer-gene maps

We downloaded per-group EpiMap enhancer-gene links from https://personal.broadinstitute.org/cboix/epimap/links/pergroup/. We lifted the genomic coordinates to GRCh38 by using LiftOver software. When we assessed aggregated EpiMap enhancer-gene links across all the 31 tissue-groups, we used “bedtools merge” function for each gene to create a union of all enhancer-gene links (**Figure 3c and d**). For tissue-specific enrichment analyses, we analyzed the 31 group-specific tracks separately (**Supplementary Figure 10a** and **10b**). To benchmark the precision and recall, we used EpiMap correlation scores to define variable sets of enhancer-gene links from EpiMap based on the threshold of EpiMap correlation score.

We downloaded ABC predictions in 131 cell types and tissues from ftp://ftp.broadinstitute.org/outgoing/lincRNA/ABC/AllPredictions.AvgHiC.ABC0.015.minus150.ForABCPaperV3.txt.gz. We lifted the genomic coordinates to GRCh38 by using LiftOver software. When we assessed aggregated ABC enhancer-gene links across all the groups, we used “bedtools merge” function for each gene to create a union of all enhancer-gene links across 131 cell types (**Figure 3c and d**). For cell-type-specific analyses, we aggregated cell lines or cell types to be corresponding with our cell types and analyzed each of these tracks separately (B cell, T cell, Myeloid cells, and fibroblasts; **Supplementary Figure 10a** and **10b**). To benchmark the precision and recall, we used ABC scores to define variable sets of enhancer-gene links from ABC model based on the threshold of ABC score.

To assess precision and recall and compare against bulk-tissue based methods (i.e., EpiMap and ABC model), we used sets of significant peak-gene linkages in each method with varying thresholds (i.e., FDR in SCENT {0.5, 0.3, 0.2, 0.1, 0.05, 0.02}, EpiMap correlation score {0, 0.4, 0.8, 0.9} in EpiMap, and ABC score {0, 0.05, 0.1, 0.2} for ABC model). We then used each set of peak-gene linkages to re-calculate causal variant enrichment for GWAS (**Figure 3d**).

We also assessed the impact of PIP threshold in defining a set of statistically fine-mapped variants on the causal variant enrichment analysis. To do so, we re-defined the set of putative causal variants with more stringent PIP thresholds (PIP > 0.5 and PIP > 0.7), and re-computed the calculate causal variant enrichment *Overall_Enrichment* score.

### caQTL analysis using scATAC-seq samples with genotype

We generated independent arthritis-tissue dataset with single-cell unimodal ATAC-seq data with genotype (*n* = 17, *manuscript in preparation*) to define chromatin accessibility QTLs (caQTLs). We used two methods, binomial test and RASQUAL. Briefly, we genotyped donors by using Illumina Multi-Ethnic Genotyping Array. We performed quality control of genotype by sample call rate > 0.99, variant call rate > 0.99, minor allele frequency > 0.01, and *P*_HWE_ > 1.0×10^−6^. We performed haplotype phasing with SHAPEIT2 software^100^ and performed whole-genome imputation by using minimac3 software^101^ with a reference panel of 1000 Genomes Project phase 3^102^. After imputation, we selected variants with imputation *Rsq* > 0.7 as post-imputation QC. We next created a merged bam file of ATAC-seq for each donor and each cell type by aggregating all the reads. Using the imputed genotype for each donor and aggregated bam files for each donor and cell type, we applied WASP^103^ to correct any bias in read mapping toward reference alleles to accurately quantify allelic imbalance. We thus created a bias-corrected bam files for each donor and cell type.

For binomial tests, we ran ASEReadCounter module in GATK software^104^ using the bias-corrected bam files as input to quantify allelic imbalance in heterozygous sites with read count > 4 within ATAC peak counts. We first performed one-sided binomial tests in each donor, and meta-analyzed the statistics across donors by Fisher’s method if multiple donors shared the same heterozygous site. For RASQUAL, we created a VCF file containing both genotype dosage and allelic imbalance from ASEReadCounter. We quantified the read coverage for each peak and for each donor by “bedtools coverage” function. We created a peak by donor matrix with read coverage. We QCed samples with log(total mapped fragments) fewer than mean – 2SD across samples in each cell type. We QCed peaks so that at least two individuals have any fragments for the peak. We then ran RASQUAL software with the inter-individual differences in ATAC peak counts (in a peak by donor matrix) and intra-individual allelic imbalance (in VCF), with accounting for chromatin accessibility PCs (the first *N* components whose explained variances are greater than those from permutation result), 3 genotype PCs, sample site and sex as covariates. RASQUAL output chi-squared statistics and *P* values. We computed FDR from these raw *P* values by Benjamini & Hochberg correction on local multiple test burden (i.e., the number of *cis*-SNPs in the region). To correct for genome-wide multiple testing, we ran the RASQUAL with random permutation, where the relationship between sample labels and the count matrix was broken. Thus, we derived *q* values for each candidate caQTL.

We finally intersected these peaks with significant caQTL effect in each significance threshold with SCENT peaks and assessed causal variants enrichment within these peaks for GWAS as explained in the previous sections.

### ClinVar analysis

We downloaded the latest clinically reported variant list registered at ClinVar from https://ftp.ncbi.nlm.nih.gov/pub/clinvar/vcf_GRCh38/clinvar.vcf.gz. We then screened the variants to exclude (1) exonic variants and (2) variants categorized as “benign”. We defined the ClinVar variant density as the number of the non-coding and non-benign variants within each annotation x 1,000 divided by the total length (bp) of each annotation.

### Somatic mutation analysis

We used a list of somatic mutation hotspot in Supplementary Table 2-20 of the original publication^95^. We lifted the genomic coordinates to GRCh38 by using LifOver software. We then intersected the non-coding somatic mutation hotspots with our cell-type-specific SCENT peaks. We compared the intersected elements’ target genes by SCENT with the “Annotate_Gene” column from the original publication.

### Downsampling experiments

To evaluate the effect of cell numbers on the statistical power in detecting significant SCENT enhancer-gene linkages, we performed downsampling experiments in fibroblast (the most abundant cell type in arthritis-tissue dataset, *n*_cell_ = 9,905). We randomly samples cells (*n*_cell_ = 500, 1000, 2500, 5000, and 7500). We then applied SCENT to each of the subset groups of cells and defined significant peak-gene links with FDR < 10%. We counted the number of significant peak-gene links in each of the subset groups of cells, and annotated peaks based on the distance to the TSS to the target gene.

## Data Availability

All data produced in the present study are available upon reasonable request to the authors

## Acknowledgments

We would like to sincerely thank participants of this study who provided tissue samples. We thank Anika Gupta, Joyce Kang and Kaitlyn Lagattuta for their comments and helpful discussion on the manuscript. This work is supported in part by funding from the National Institutes of Health (R01AR063759, U01HG012009, UC2AR081023). S.S. was in part supported by the Uehara Memorial Foundation and The Osamu Hayaishi Memorial Scholarship. K.Wei is supported by a Burroughs Wellcome Fund Career Awards for Medical Scientists, a Doris Duke Charitable Foundation Clinical Scientist Development Award, and a Rheumatology Research Foundation Innovative Research Award. We would like to thank the Brigham and Women’s Hospital Center for Cellular Profiling Single Cell Mulitomics Core for experimental design and protocol optimization.

## Author Contributions

S.S. and S.R. conceived the work and wrote the manuscript with critical input from co-authors. S.S. and K. Weinand analyzed the arthritis-tissue dataset and S.S. analyzed publicly available datasets with help and guidance from K.K.D., K.J., M.K., A.M., A.L.P., and S.R. G.F.M.W., Z.Z., M.B.B., L.T.D., and K.Wei provided samples and generated the arthritis-tissue dataset. S.I. refactored the SCENT software implementation as an R package.

## Competing Financial Interests

We declare no conflict of interest for this study. S.R. is a founder for Mestag, Inc, a scientific advisor for Rheos, Jannsen, and Pfizer, and serves as a consultant for Sanofi and Abbvie.

## Notes

### Competing Interest Statement

The authors have declared no competing interest.

### Author Declarations

Study used samples collected underIRB approved protocols at Brigham and Women's Hospital and Hospital for Special Surgery.

### Summary of Updates

Additional analyses and clarifications with additional supplementary figures

