## Supplementary Figures for "Tissue-specific enhancer-gene maps from multimodal single-cell data identify causal disease alleles"

**Sakaue et al.**

**Table of contents:**

Page 3 **Supplementary Figure 1**

Page 4 **Supplementary Figure 2**

Page 5 **Supplementary Figure 3**

Page 6 **Supplementary Figure 4**

Page 7 **Supplementary Figure 5**

Page 8 **Supplementary Figure 6**

Page 9 **Supplementary Figure 7**

Page 10 **Supplementary Figure 8**

Page 11 **Supplementary Figure 9**

Page 12 **Supplementary Figure 10**

Page 14 **Supplementary Figure 11**

Page 15 **Supplementary Figure 12**

Page 16 **Supplementary Figure 13**

Page 17 **Supplementary Figure 14**

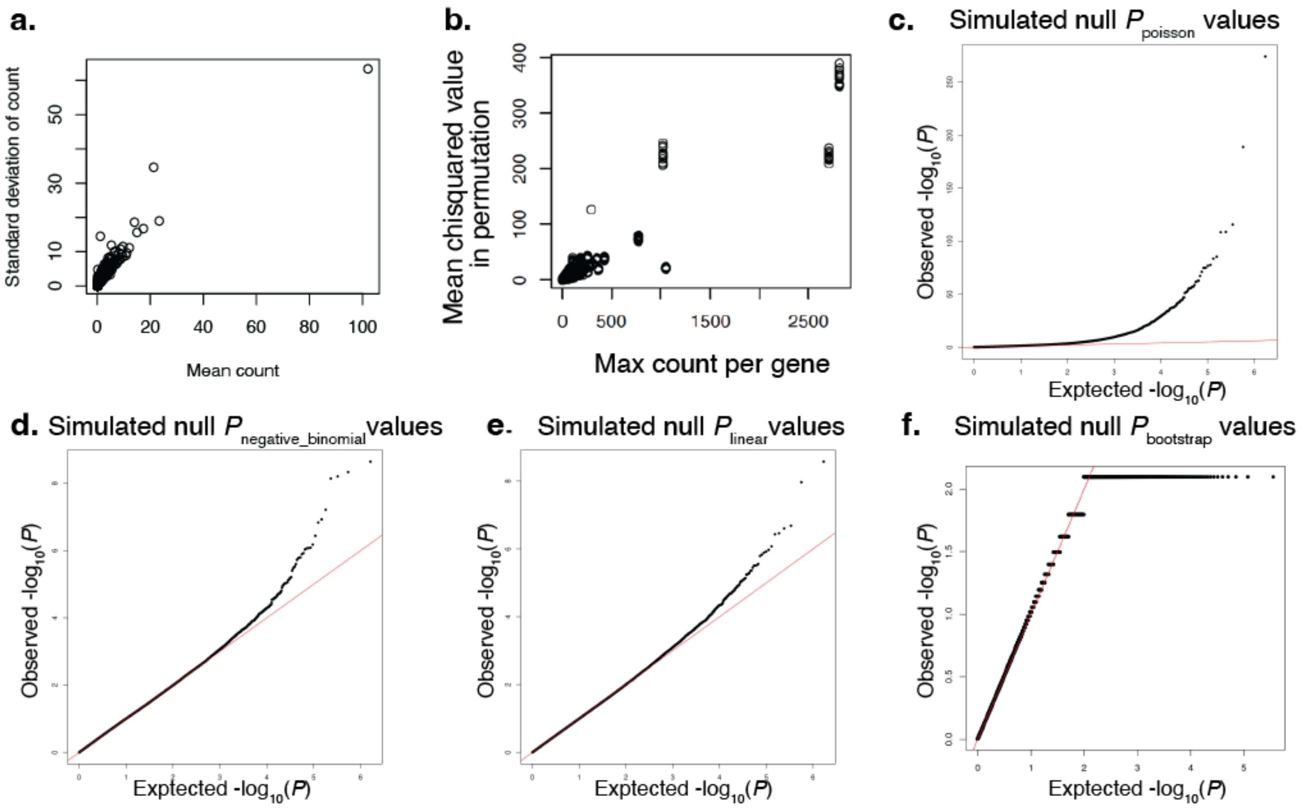


**Supplementary Figure 1 | Distribution of gene expression counts in single-cell RNA-seq and statistics from association between gene expression and chromatin accessibility under null simulation.**

**a**. In an example dataset of arthritis-dataset, mean gene count was strongly correlated with standard deviation of the gene count. **b**. The correlation between max expression count per gene (x-axis) and the mean naïve association chi-square values ($\chi^2$) from Poisson regression between gene expression and chromatin accessibility under null simulation (y-axis). **c**. The quantile-quantile (QQ) plot of *P* values from the Poisson regression between gene expression count and chromatin accessibility under null simulation. **d**. The QQ plot of *P* values from the negative binomial regression between gene expression count and chromatin accessibility under null simulation. **e**. The QQ plot of *P* values from the linear regression between log-normalized and inverse-normal-transformed gene expression and chromatin accessibility under null simulation. **f**. The QQ plot of *P* values estimated from bootstrapping based on the statistics distributions from the Poisson regression between gene expression count and chromatin accessibility under null simulation.

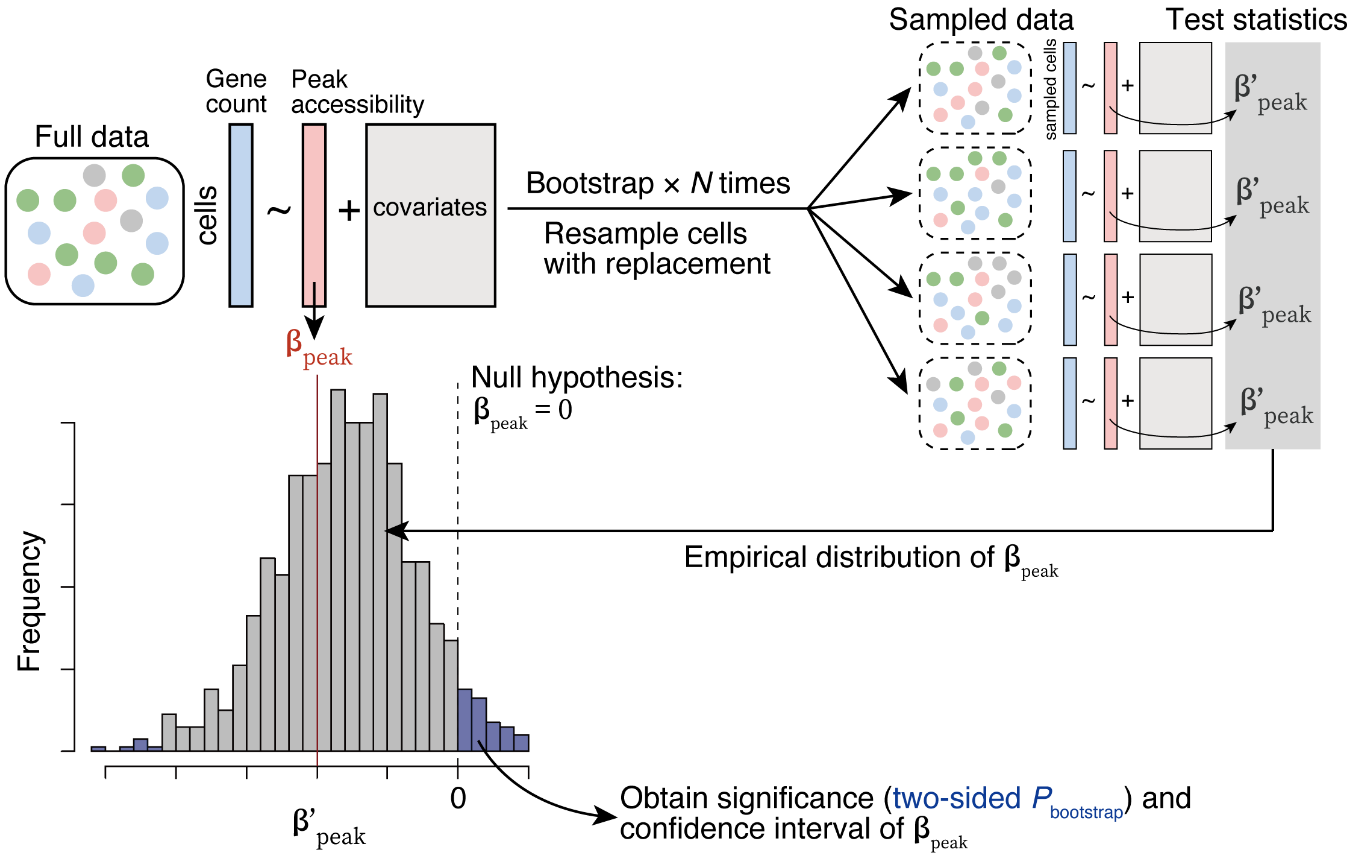


**Supplementary Figure 2 | Schematic overview of SCENT model using Poisson regression and non-parametric bootstrapping.**

We first run Poisson regression associating the raw gene expression count (RNA-seq) with the peak accessibility (ATAC-seq) accounting for technical covariates across the entire cells in the multimodal data to estimate $\beta_{peak}$. Then, we resampled cells with replacement from the full data in each of the bootstrapping round and re-estimated $\beta'_{peak}$ for *N* times. We compared this empirical distribution of $\beta'_{peak}$ against the null hypothesis ($\beta'_{peak}$ = 0) to derive the significance of $\beta_{peak}$ (i.e., two-sided bootstrapping-based *P* value = $P_{\text{bootstrap}}$).

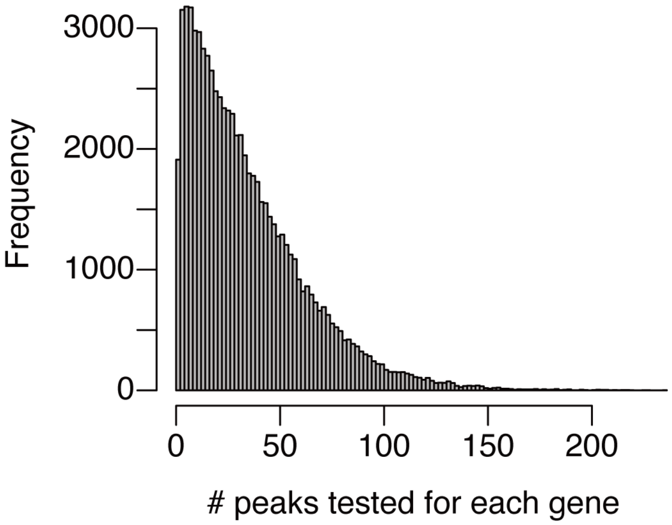


**Supplementary Figure 3 | The number of ATAC-seq peaks for each gene for SCENT association tests.**

In each dataset, we only performed association tests between genes and ATAC-seq peaks that are in *cis* regions from the gene body (< 500 kb). Thus, each gene has different number of ATAC-seq peaks tested for SCENT associations. This histogram shows the mean number of ATAC-seq peaks tested for each gene (i.e., *cis*-peaks from the gene) across all the datasets.

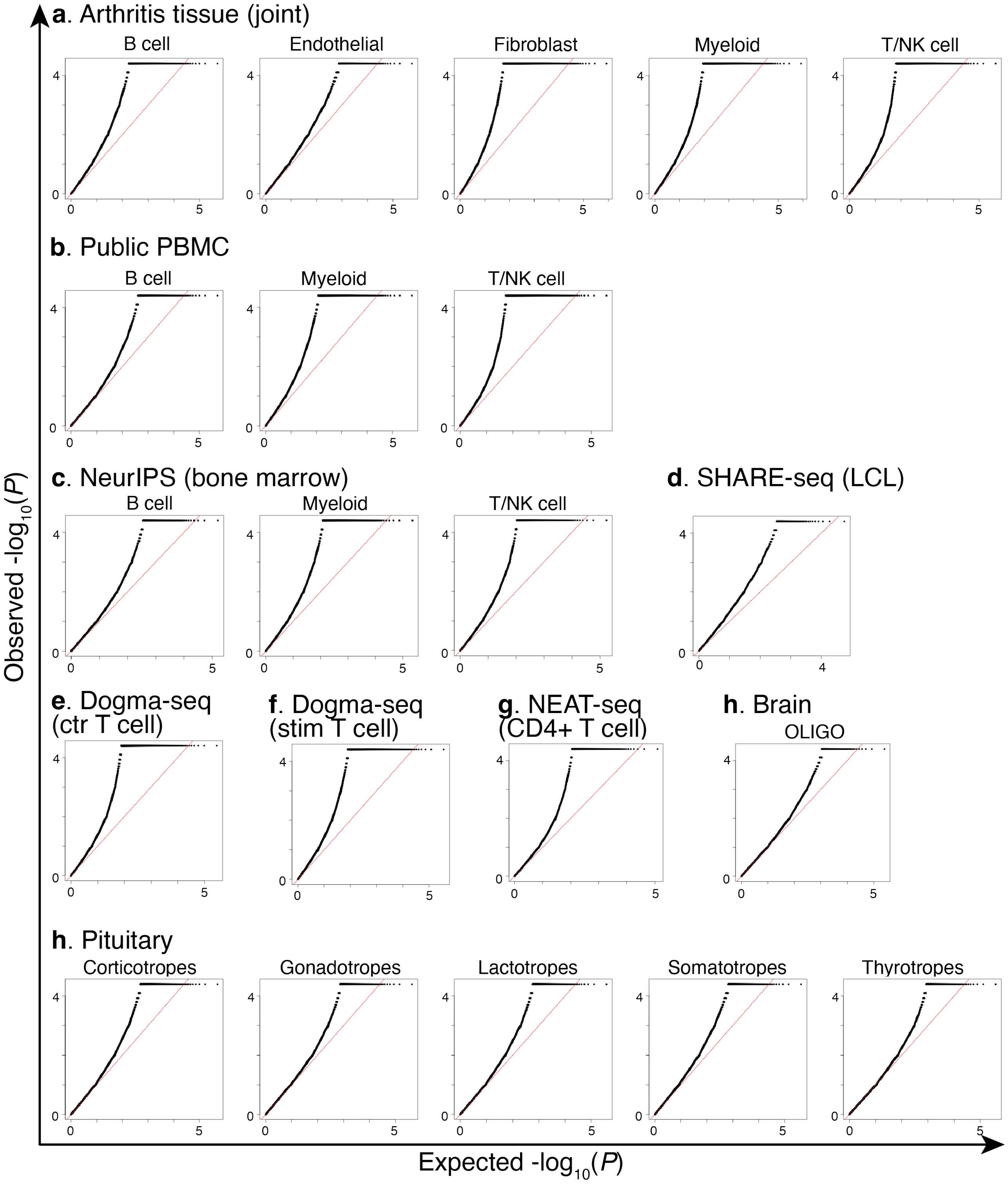


**Supplementary Figure 4 | The QQ plot of SCENT P values by bootstrapping.**

We applied SCENT to each of 23 broad cell types from 9 single-cell multimodal datasets. Each QQ plot represents $P_{bootstrap}$ values in each cell type in each dataset.

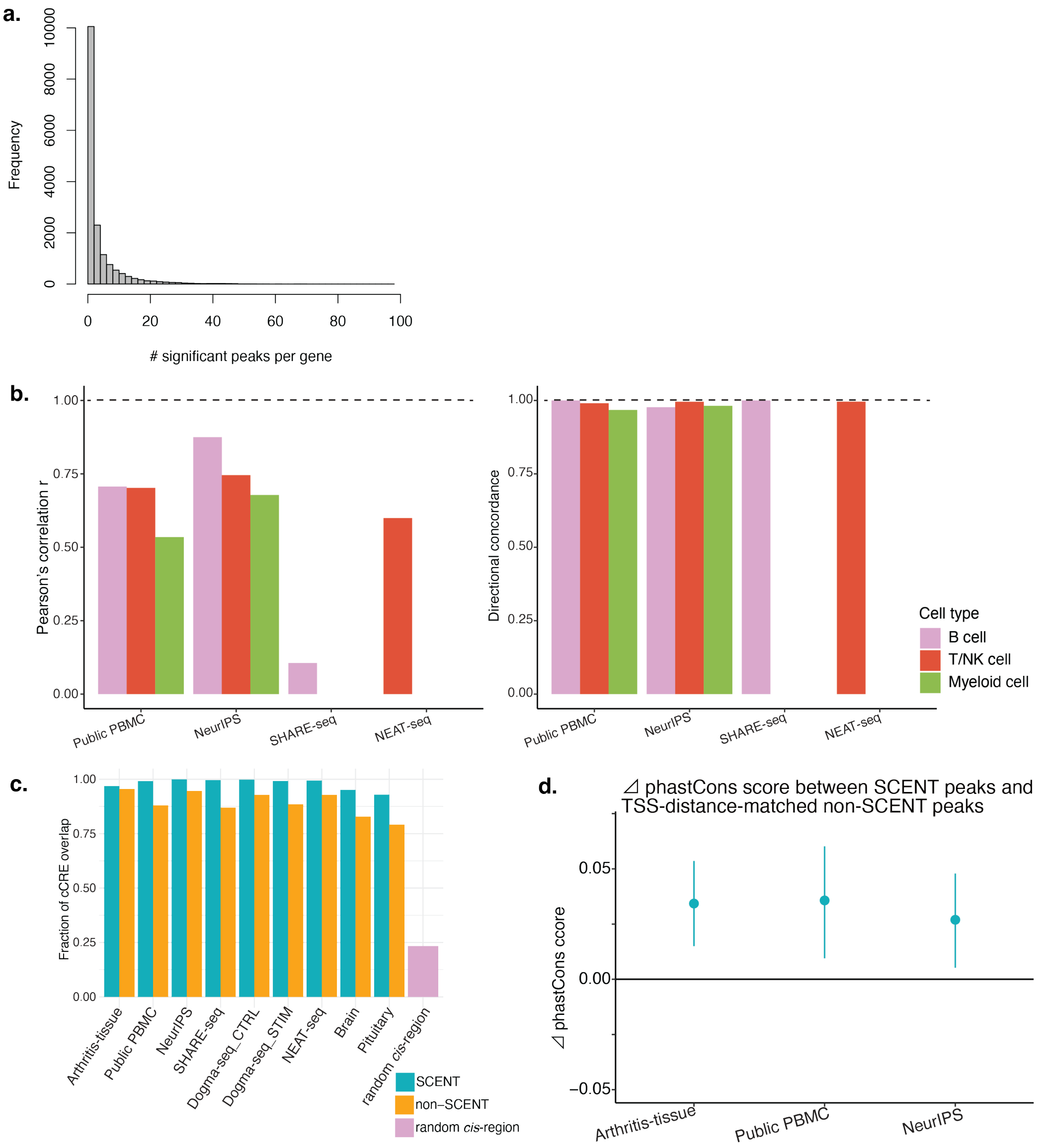


**Supplementary Figure 5 | Properties of SCENT peaks.**

**a**. The number of significant SCENT peaks per gene across genes we investigated in at least one dataset-cell type pair. **b**. The effect size correlation *r* by Pearson's correlation between arthritis-tissue dataset and the other dataset for the same cell type (left) and the directional (sign) concordance between arthritis-tissue dataset and the other dataset for the same cell type (right). **c**. Fraction of overlap with ENCODE cCREs in SCENT (green) or non-SCENT peaks (orange) in each dataset and random set of *cis*-non-coding regions (pink). **d**. The mean Δ phastCons score between SCENT peaks and TSS-distance-matched non-SCENT peaks across all the genes. The bars indicate the 95% CI by bootstrapping genes.

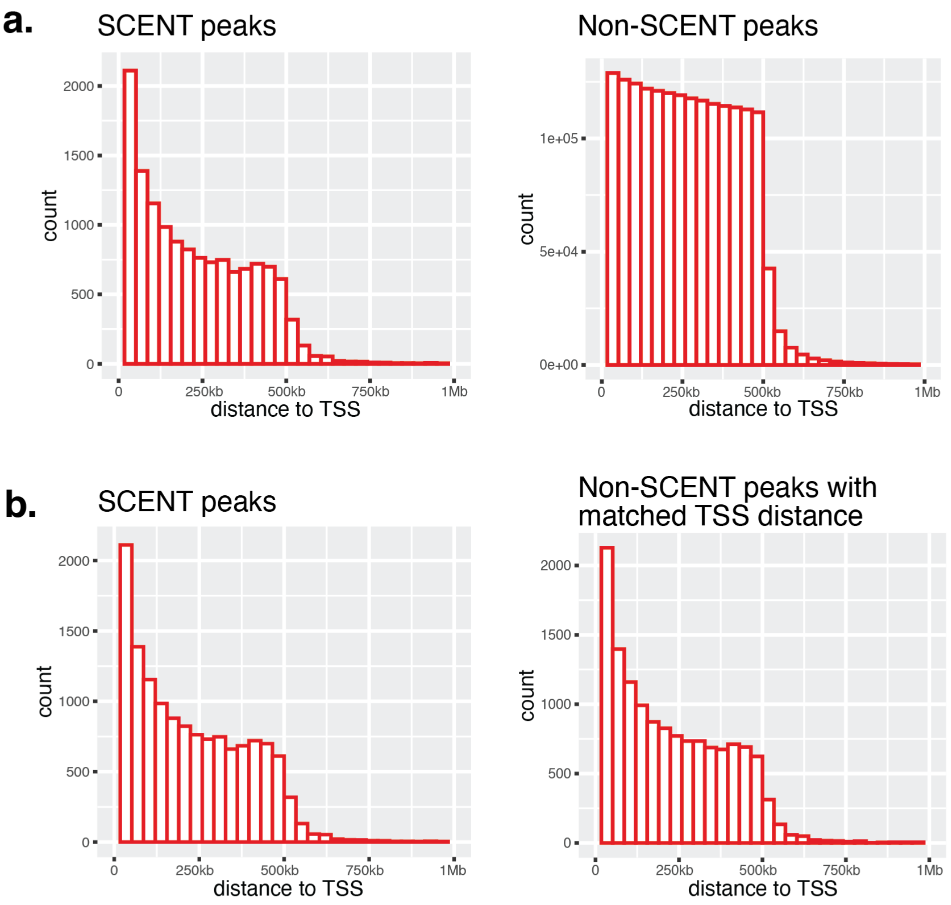


**Supplementary Figure 6 | The distance between the SCENT/non-SCENT peaks and the transcription start site (TSS) of the target gene.**

**a.** Shown are histograms of SCENT or non-SCENT peaks' distance to the transcription start site (TSS). **b**. Histograms of TSS-distance-matched SCENT or non-SCENT peaks' distance to the TSS (see **Methods**).

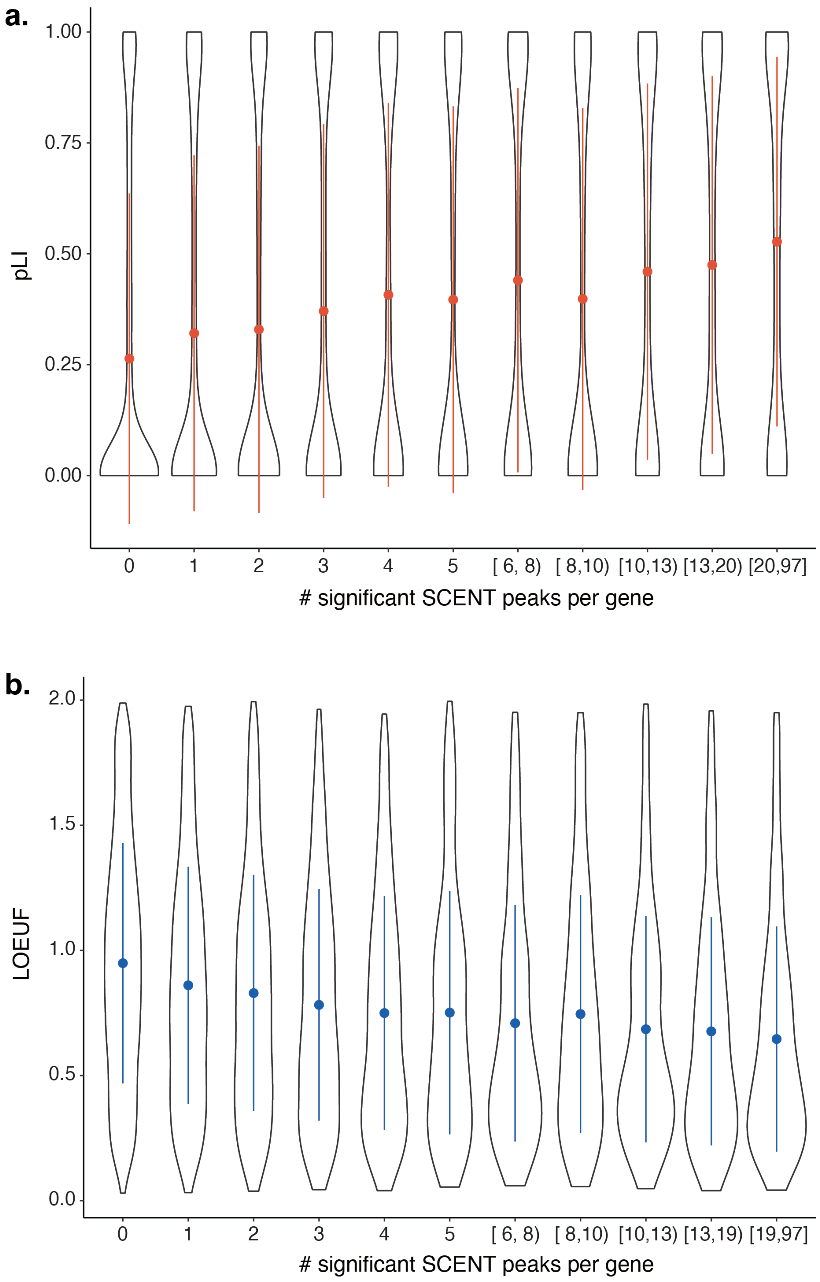


**Supplementary Figure 7 | Mutational constraint on genes with a high number of SCENT peaks.**

For each gene, the number of SCENT peaks were counted and binned as shown in the x-axis, and mutational constraint metric (pLI: **a**, LOEUF: **b**) for genes within each bin are shown as a violin plot on the y-axis.

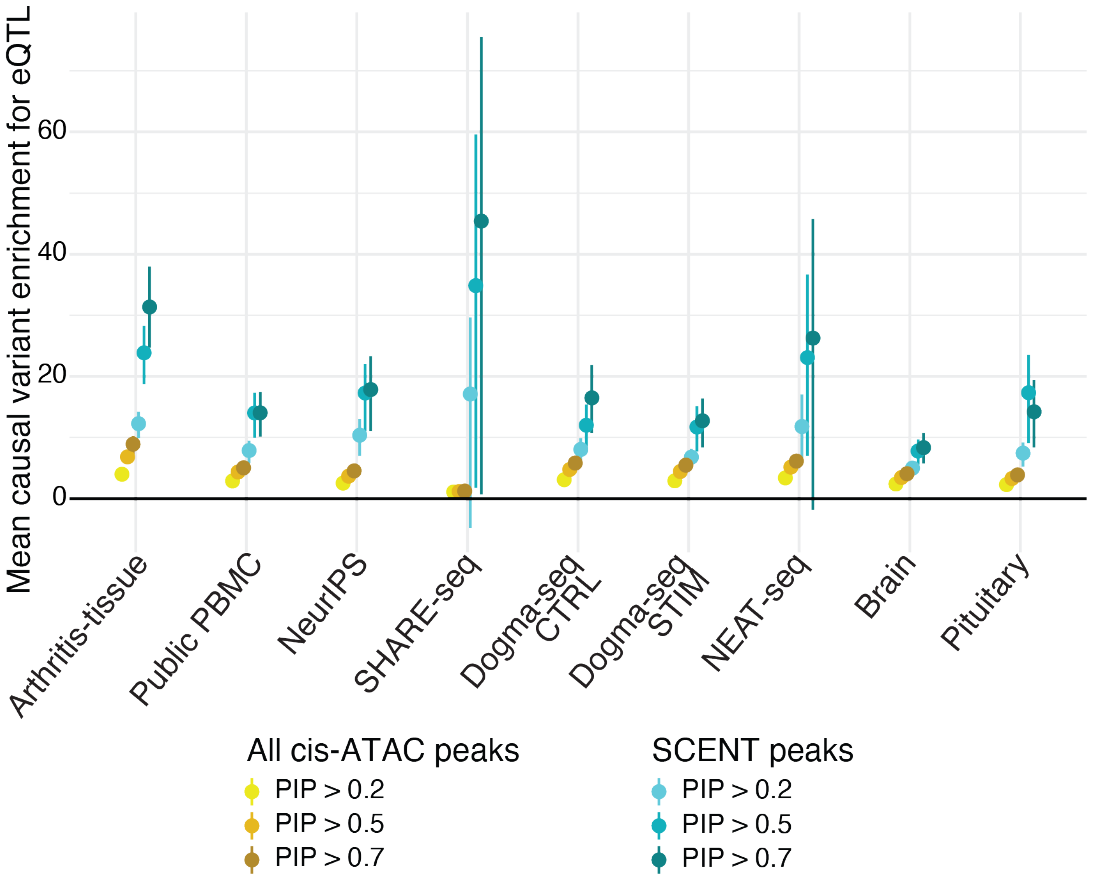


**Supplementary Figure 8 | Causal variant enrichment for eQTL in each of the PIP threshold.**

We calculated the causal variant enrichment in all cis-ATAC peaks and SCENT peaks in each dataset by changing the PIP thresholds {>0.2,>0.5,>0.7} in defining putative causal variants from fine-mapping.

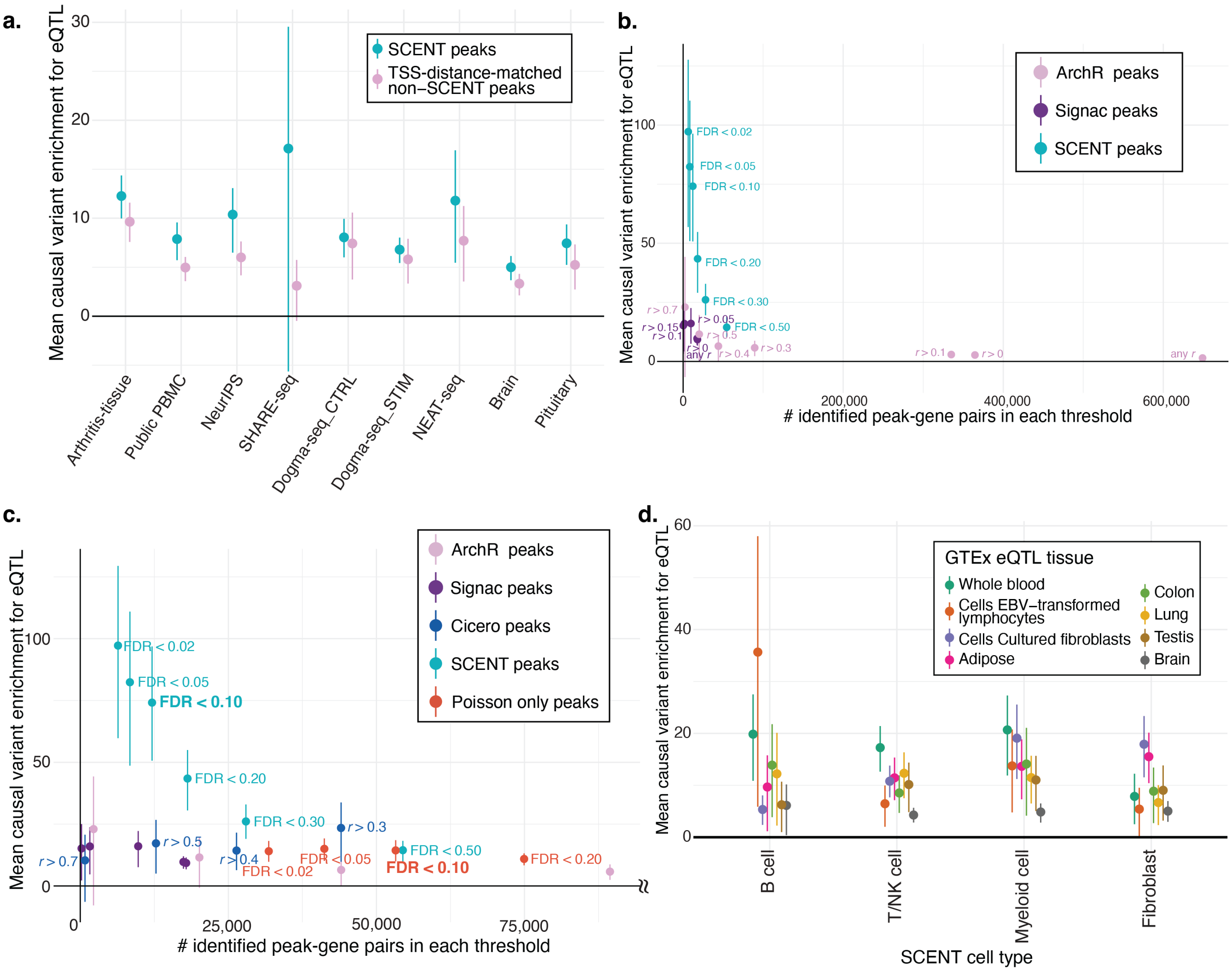


**Supplementary Figure 9 | Causal variant enrichment for eQTLs.**

**a**. The mean causal variant enrichment for eQTL variants within SCENT peaks (green) or non-SCENT peaks with matching disctance to TSS (pink). **b**. Comparison of the mean causal variant enrichment for eQTL (y-axis) among SCENT (green), ArchR (pink), and Signac (purple) as a function of the number of significant peak-gene pairs at each threshold of significance. **c**. Comparison of the mean causal variant enrichment for eQTL (y-axis) among original SCENT (Poisson regression + non-parametric bootstrapping; green), Poisson-only strategy without bootstrapping (red), ArchR (pink), Signac (purple), and Cicero (correlation method using sc-ATAC-seq alone; blue) as a function of the number of significant peak-gene pairs at each threshold of significance up to 100,000 peak-gene linkages. **d**. Tissue-specific causal variant enrichment within SCENT peaks. The dots and lines are colored by the eQTL source tissue in GTEx that we assessed. Lines indicate 95% confidence interval by bootstrapping. In all panels, the bars indicate 95% confidence intervals by bootstrapping genes.

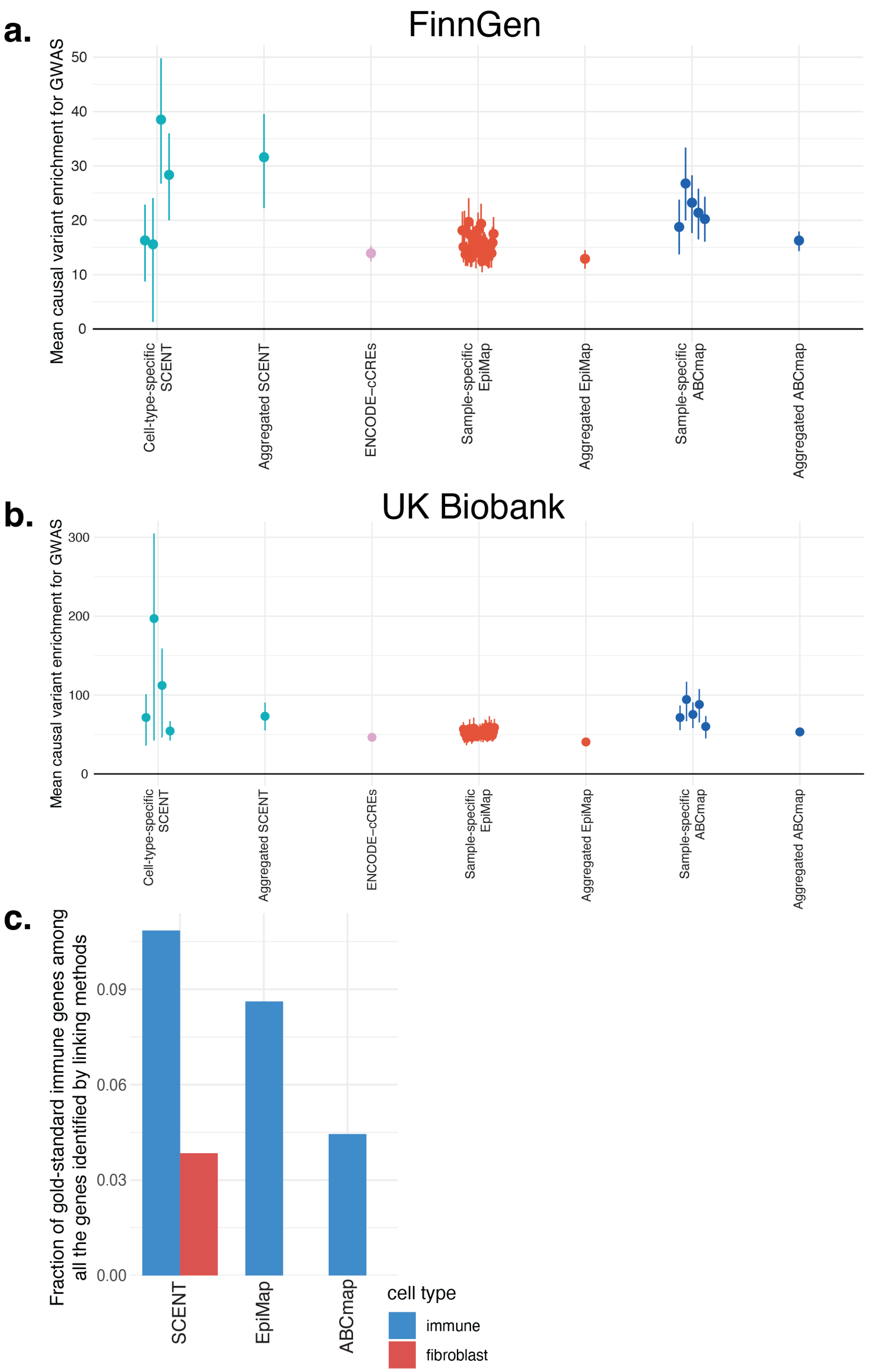


**Supplementary Figure 10 | Causal variant enrichment for GWAS**

**a** and **b**. The mean causal variant enrichment for GWAS within cell-type-specific and aggregated SCENT enhancers (green), ENCODE cCREs (pink), group-specific and aggregated EpiMap enhancers (red) and sample-specific and aggregated ABC enhancers (blue). GWAS results were based on FinnGen (**a**) and UK Biobank (**b**). The bars indicate 95% confidence intervals by bootstrapping traits. **c**. The fraction of known genes from Mendelian autoimmune diseases among all the genes identified by SCENT, EpiMap, and ABC model. The color of the bars indicates the cell types in each linking method.

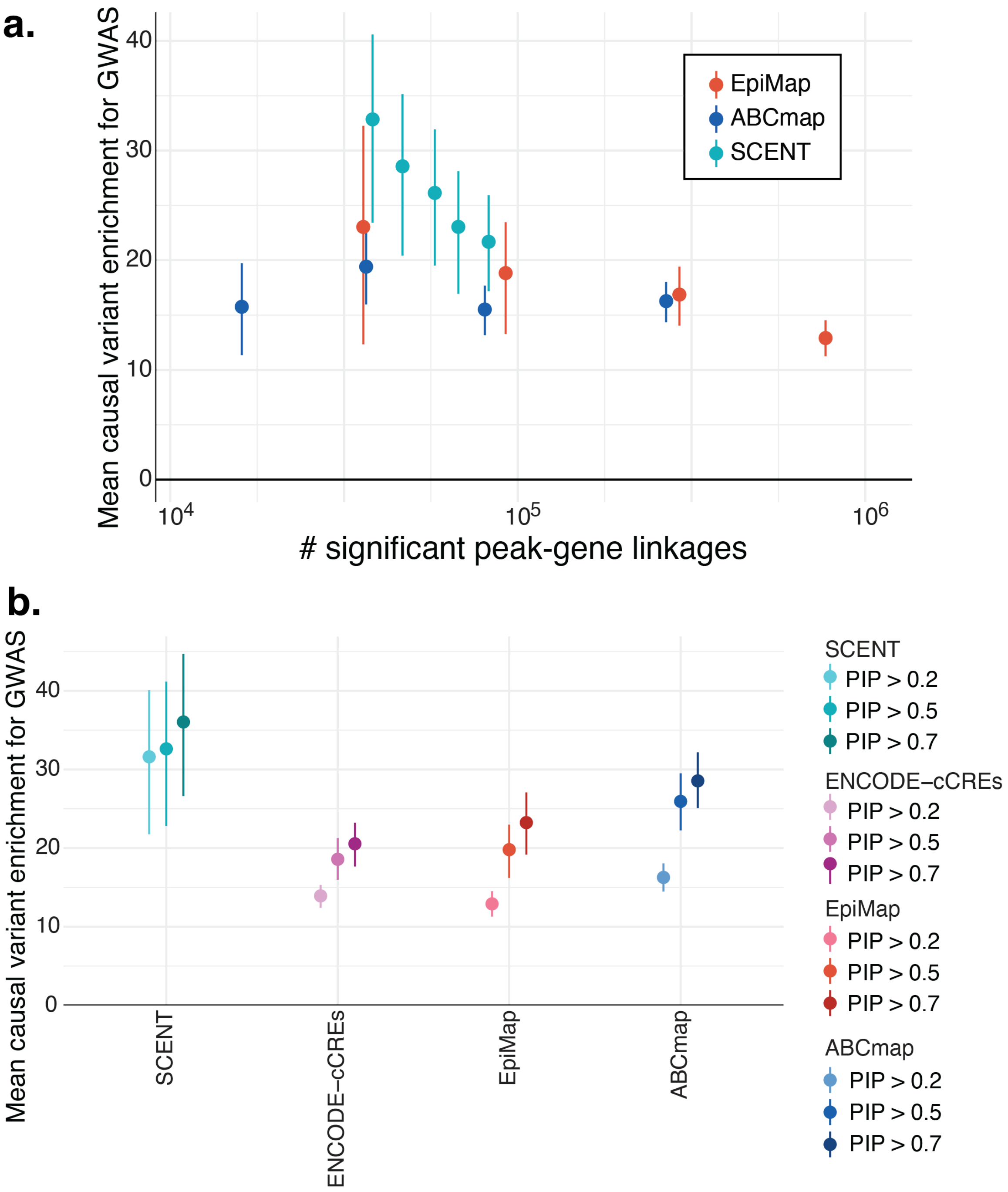


**Supplementary Figure 11 | Causal variant enrichment for GWAS and comparison with EpiMap and ABC model**

**a**. Comparison of the mean causal variant enrichment for FinnGen GWAS (y-axis) among SCENT (green), EpiMap (red), and ABC model (blue) as a function of the number of significant peak-gene pairs (x-axis) at each threshold of significance. The bars indicate 95% confidence intervals by bootstrapping traits. **b**. We calculated the causal variant enrichment for FinnGen GWAS among SCENT (greens), EpiMap (reds), and ABC model (blues) by changing the PIP thresholds in defining putative causal variants from fine-mapping.

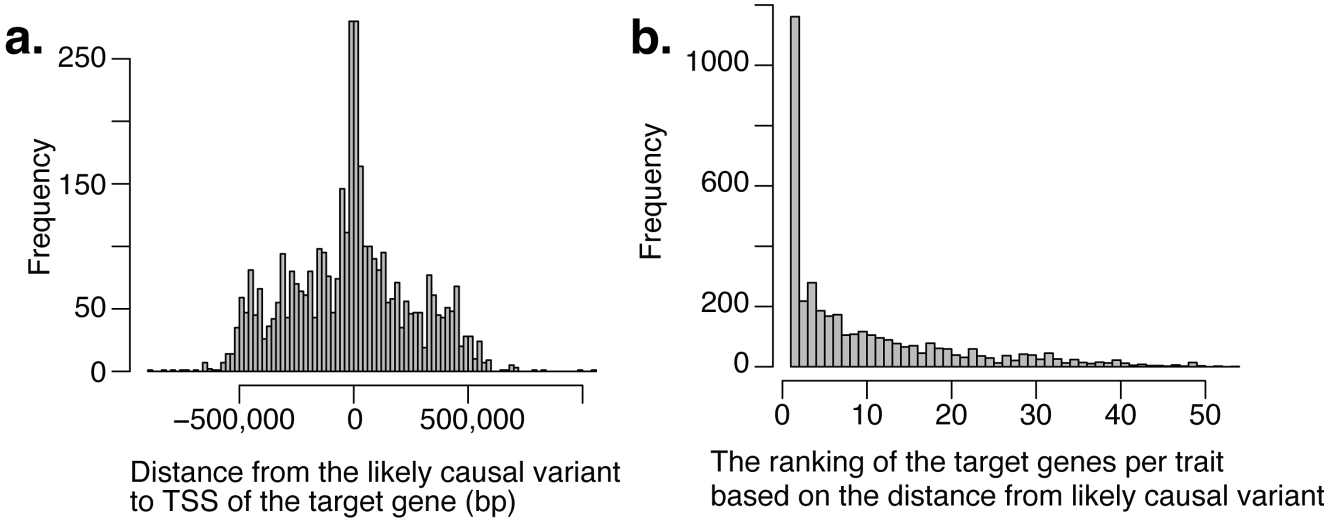


**Supplementary Figure 12 | The putative causal genes for potential GWAS causal variants.**
**a**. Shown is a histogram of the distance in bp between the putative causal variants from GWAS and the transcription start site (TSS) of their corresponding target genes identified by SCENT. **b**. The rank of the identified target genes for each trait by SCENT when we ordered all the genes based on the distance between the putative causal variant and the TSS of the genes.

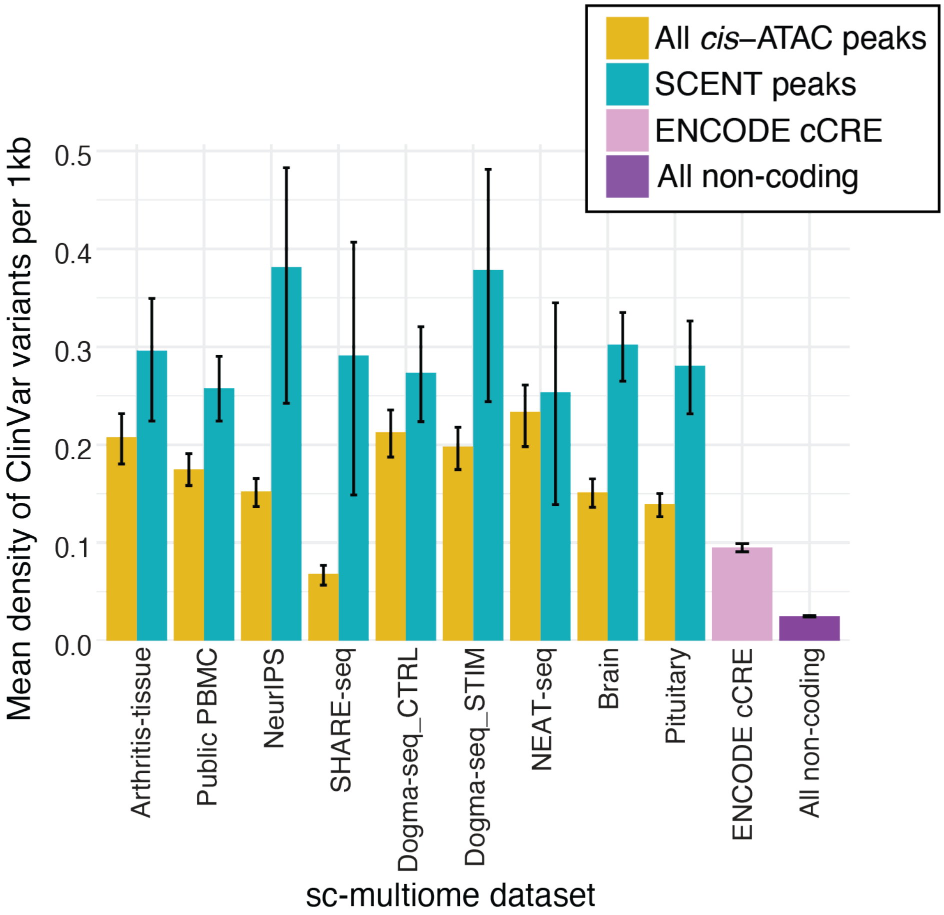


**Supplementary Figure 13 | ClinVar density comparison.**

The mean number of ClinVar non-benign non-coding variants per 1 kb within SCENT peaks, all *cis*-ATAC peaks, ENCODE cCREs, and all non-coding *cis*-regions from genes. The bars indicate the 95% CI by bootstrapping the regions in each category.

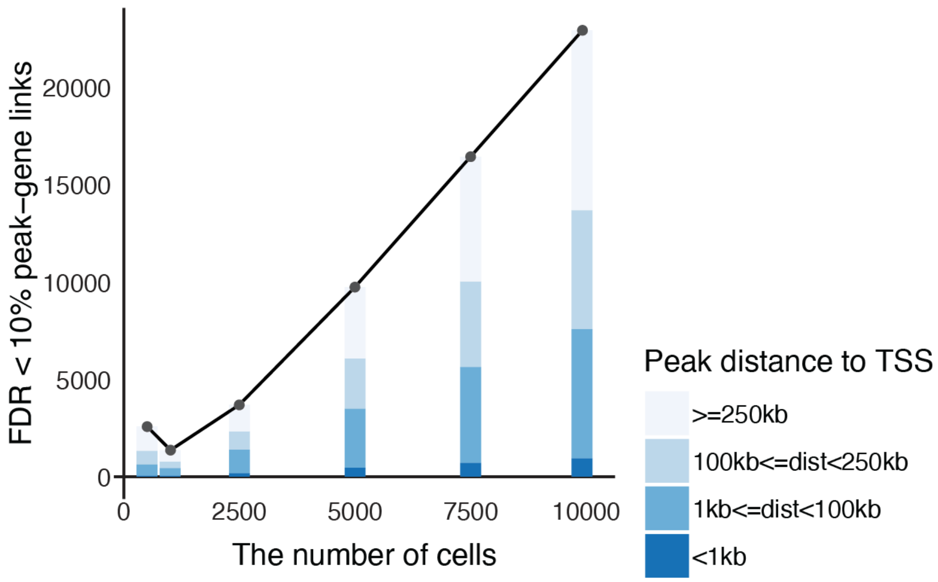


**Supplementary Figure 14 | Downsampling experiment of SCENT.**

We downsampled cells in arthritis-tissue dataset (fibroblasts) and reran SCENT for those cells to define significant peak-gene linkages. The x-axis represents the number of cells and the y-axis represents the number of FDR-significant peak-gene linkages in each downsampling experiment. The stacked bars indicates peaks' distance with regard to the TSS of the target gene.
